# Exploring the Application of the Observational Medical Outcomes Partnership Common Data Model to Multi-site Stroke Rehabilitation Research Data

**DOI:** 10.64898/2026.06.28.26356618

**Authors:** Katherine J. Loomis, Amisha Kumar, Octavio Marin-Pardo, Grace C. Bellinger, Margaret A. French, Ryan T. Roemmich, Sook-Lei Liew

## Abstract

**Background:** Emerging artificial intelligence and machine learning (AI/ML) tools can help generate robust knowledge to support precision rehabilitation approaches for varied patient populations. There is a large amount of research-generated and clinical rehabilitation data available for this purpose; however, a pronounced lack of interoperability prevents large-scale data aggregation. Common data models (CDMs) such as Observational Medical Outcomes Partnership (OMOP) have improved data interoperability across healthcare settings, and more recently, for clinical rehabilitation data, specifically. However, the application of these CDMs to research-generated data has not yet been explored. Therefore, as a foundational step, our study evaluated the breadth and depth of OMOP CDM coverage for data in a multi-site repository of harmonized rehabilitation research data: the Enhancing NeuroImaging Genetics through Meta-Analysis Stroke Recovery (ENIGMA-SR) database.

**Methods:** Two raters independently mapped data elements representing 46 demographics and medical history (DMH) ENIGMA-SR variables and 95 distinct ENIGMA-SR rehabilitation assessments to OMOP standard concepts. Initial rater agreement was assessed for data element inclusion in OMOP and for specific OMOP concepts used (primary metric: Gwet’s agreement coefficient [AC]). Mapping differences were reconciled, and final mappings were descriptively analyzed to examine (1) overall OMOP inclusion, (2) inclusion of more granular levels (subscales, items) of complex assessments, and (3) mapped OMOP concept characteristics.

**Results:** Initial rater agreement was good/very good for overall OMOP inclusion of DMH and assessment data elements and for OMOP concepts mapped across almost all assessment data elements (Gwet’s AC: 0.79-0.89). Initial OMOP concept agreement was more variable for DMH data elements; however, all mapping differences were successfully reconciled to 100%. Overall, DMH data elements had higher OMOP inclusion than rehabilitation assessments: 84.8% (39/46) vs. 58.9% (56/95). OMOP coverage was particularly limited for complex assessment subscale- and item-level data elements (9.4% [3/32]; 19.2% [14/73]) and did not match the granularity level represented in ENIGMA-SR data for 56.2% (41/73) of complex assessments. DMH and top-level assessment data elements were frequently mapped to multiple OMOP concepts (median: 6, 2; range: 1-23, 1-8), and for > 50% of these data elements the concepts spanned 2-3 different OMOP domains.

**Conclusion:** For ENIGMA-SR, the OMOP CDM has good coverage of DMH data, moderate top-level coverage of rehabilitation assessments, and very limited coverage of assessment subscales and items. This uneven coverage, combined with variability in OMOP concepts and domains mapped to equivalent data points, presents challenges for aggregating clinical and research-generated rehabilitation data into AI/ML-ready datasets. Moreover, software tools currently available to facilitate the mapping process do not effectively accommodate content- and structure-related features inherent to research-generated data. Going forward, the utility of the OMOP CDM to aggregate multi-source rehabilitation data may be improved by expanding the catalogue of OMOP rehabilitation-related concepts, building cross-walks to research-oriented data standards, and adapting emerging computational tools to streamline the mapping process.

## 1. Background and Purpose

Increasingly rapid innovations in the fields of artificial intelligence (AI) and machine learning (ML) provide incredible opportunities to harness powerful analytical tools to advance foundational knowledge needed for the development of precision rehabilitation approaches.^1^ Ultimately, these approaches can help rehabilitation service providers better address the needs of diverse patient populations and improve the overall efficiency of rehabilitative care. Simultaneously, the widespread adoption of electronic health records (EHRs),^2-4^ as well as initiatives such as the revised NIH Policy on Data Management and Sharing and other funding and academic journal mandates^5,6^ have produced a tremendous quantity of digitally accessible clinical and research data. Successful aggregation of these existing datasets could result in large-scale, diverse AI/ML-ready datasets, reducing the burden on individuals to independently construct their own large datasets for AI/ML training.

The greatest barrier to aggregation of both EHR and research-generated rehabilitation data is the lack of data interoperability that results from the siloed nature of data collection. Interoperability here refers to the ability of data to be integrated across different systems and contexts while retaining equivalent meaning. Different EHR systems, sites, studies, and databases use variable (and often incompatible) data terminology, structure, and attributes, even when referring to equivalent constructs or assessments.^4,7-11^ For EHR data specifically, as emerging automated approaches enable more reliable extraction of structured data from unstructured text (e.g., clinical notes), interoperability challenges may be compounded over time, since this data has the potential for even greater variability.^4,12-14^ For research-generated rehabilitation data, additional variability results from the prevalence of small-scale, independently conducted studies generating data within limited conceptual scopes and the absence of widely adopted standards governing data content or form.

Over the past several decades, common data models (CDMs; e.g., Observational Medical Outcomes Partnership [OMOP], Sentinel, the National Patient-Centered Clinical Research Network [PCORnet], and Informatics for Integrating Biology in the Bedside)^15,16^ have been developed that specify standard data terms and structure to improve the interoperability of EHR data from different medical contexts. More recently, there has been growing advocacy specifically for more sophisticated and comprehensive CDM inclusion of rehabilitation-related concepts and for the expanded use of CDMs to facilitate successful aggregation of rehabilitation EHR data.^12,16,17^ However, these efforts have not yet extended to research-generated rehabilitation data. Therefore, it is pertinent to evaluate the ability of clinical CDMs to (1) serve as a bridge between clinical and research-generated rehabilitation data, (2) unlock access for research data to established robust data harmonization and analytic pipelines, and (3) enhance the diversity and volume of rehabilitation datasets to be AI/ML-ready. Combined clinical- and research-generated datasets can further enable analyses comparing laboratory versus real-world practice population characteristics and outcomes, offering insight into the optimal translation of interventions from research to practice.

As a foundational step, we will explore the ability of one of the most widely used and well-supported open-source clinical CDMs, OMOP,^18-20^ to represent data within a harmonized large-scale, multi-site rehabilitation research data repository: the Enhancing NeuroImaging Genetics through Meta-Analysis Stroke Recovery (ENIGMA-SR) database.^21^ Over the past 10 years, the ENIGMA-SR database has evolved from collecting research-generated neuroimaging data, demographic data, and a single motor function outcome, to collecting various outcomes across diverse categories related to rehabilitation. To date, it contains data from almost 2,400 participants across 62 research cohorts from 14 countries around the world, providing a diverse assortment of research-based rehabilitation variables for which OMOP coverage can be examined. The purpose of this project is to characterize the breadth and depth at which variables from a large-scale stroke rehabilitation research database are covered within the OMOP CDM. In doing so we aimed to (1) evaluate the utility of the OMOP CDM to facilitate the aggregation of large-scale rehabilitation research data with clinical data and (2) identify gaps to inform targeted expansion of OMOP CDM or the development of supplementary data schemas, tools, or processes.

## 2. Methods

This study aimed to map rehabilitation-related data elements from the ENIGMA-SR database to OMOP concepts, following a vetted multi-step process of: (1) generating and categorizing ‘proxy’ data elements to represent the source data, (2) performing initial mapping via two raters, (3) reconciling mapping differences between raters, and (4) producing a final mapping list.^17^ The final list was then analyzed descriptively to characterize the breadth and depth of OMOP coverage of database variables.

### 2.1. Source Data

For the purposes of the current study, we evaluated OMOP coverage specifically of non-imaging (rehabilitation) variables (n = 932) in the ENIGMA-SR database across two broad categories: (1) demographics and medical history (DMH) and (2) assessments.

ENIGMA-SR DMH variables (n = 46) include data related to: (1) demographics, (2) stroke-related medical history, (3) other medical history, and (4) care/study-related information (**Table 1**). Almost half of the DMH variables represent stroke-related constructs (stroke-related conditions, or information on stroke recovery). Rehabilitation assessment variables (n = 886) include data from 95 distinct assessments with data at various levels of granularity across the categories of: (1) cognitive, (2) global function, (3) psychosocial, and (4) sensorimotor (**Table 2**). Only five assessments are stroke-specific (developed specifically for patients recovering from stroke). To improve the interpretability of mapping results and enable uniform, structured analyses, we represented real-world ENIGMA-SR variables with ‘proxy’ data elements. Each DMH variable was represented 1-to-1 by a single data element, while assessment variables were collapsed into a single data element per distinct assessment (removing laterality and anatomical region) to avoid over-representation. For complex assessments, defined as having multiple items and/or generating multiple data points (n = 73; e.g., the Action Research Arm Test [ARAT] or multi-trial assessments such as Stroop Color and Word Test), we added one more data element representing item-level data (n = 73), and for those having formal subscales, we added a third data element representing subscale-level data (n = 32). For example, the ARAT was represented by three data elements: (1) overall assessment, (2) subscales (grasp, grip, pinch, gross), and (3) items. The presence of multiple items/data points and subscales was determined via assessment-related publications, publicly available assessment manuals, and the Shirley Ryan Ability-Lab Rehabilitation Measures Database,^22^ and associated proxy data elements were generated regardless of whether ENIGMA-SR included each specific level of assessment data. These additions resulted in 200 total assessment data elements for a grand total of 246 data elements overall (DMH + assessment).

**Table 1.**
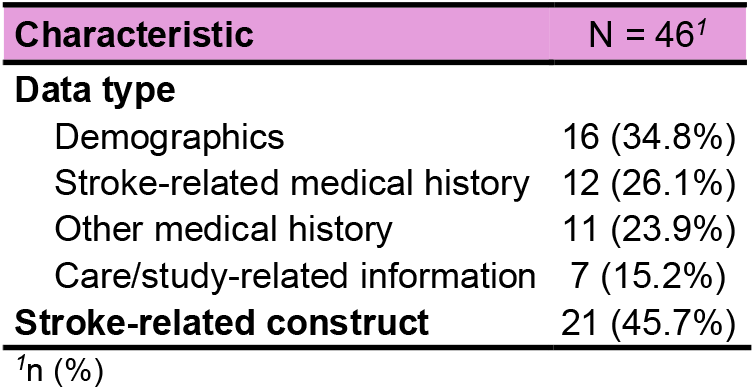
ENIGMA-SR Demographics and medical history variable characteristics.

| Characteristic | N = 46<sup>1</sup> |
| --- | --- |
| **Data type** |  |
| Demographics | 16 (34.8%) |
| Stroke-related medical history | 12 (26.1%) |
| Other medical history | 11 (23.9%) |
| Care/study-related information | 7 (15.2%) |
| **Stroke-related construct** | 21 (45.7%) |

<sup>1</sup>n (%)

**Table 2.** ENIGMA-SR Assessment characteristics.

| Characteristic | All Assessments<br>N = 95<sup>1</sup> | Complex Assessments<br>N = 73<sup>1,2</sup> | Complex Assessments with Subscales<br>N = 32<sup>1,2</sup> |
| --- | --- | --- | --- |
| **Assessment type** |  |  |  |
| Cognitive | 33 (34.7%) | 28 (38.4%) | 12 (37.5%) |
| Global function | 16 (16.8%) | 14 (19.2%) | 7 (21.9%) |
| Psychosocial | 13 (13.7%) | 13 (17.8%) | 5 (15.6%) |
| Sensorimotor | 33 (34.7%) | 18 (24.7%) | 8 (25.0%) |
| **Stroke-specific assessment** | 5 (5.3%) | 5 (6.8%) | 3 (9.4%) |

<sup>1</sup>n (%)
<sup>2</sup>Complex assessment is defined as an assessment that has more than one item and/or produces more than one datapoint

### 2.2. Initial Mapping Procedures

Two raters with extensive knowledge of ENIGMA-SR database variables independently attempted to map all 246 data elements to OMOP concepts using two open-source tools developed by Observational Health Data Sciences and Informatics (OHDSI) collaborative to facilitate the use of the OMOP CDM: (1) USAGI^23^ as the primary tool and (2) Automated Terminology Harmonization, Extraction and Normalization for Analytics (Athena)^24^ as a supplemental tool when needed. USAGI uses a term similarity algorithm to generate automated OMOP concept mapping suggestions and was used as the primary mapping tool. Athena is a web-based search engine used to browse OMOP concepts and was used to provide supplemental information to aid mapping decisions. A protocol was also developed to standardize mapping procedures between raters (**Supplemental Document 1**). Mapping was performed exhaustively for all data elements, where all directly related OMOP concepts were mapped to each data element (rather than 1 OMOP concept per element), and OMOP concept selection was limited to standard (rather than non-standard) OMOP concepts. Initial mapping results for each rater were exported from USAGI for analysis and reconciliation.

Raters reviewed the USAGI OMOP concept suggestions for each DMH and assessment data element to determine whether they were appropriate and whether there were additional OMOP concepts to which data elements were appropriate to map. For complex assessment subscale- and item-level data elements, raters also examined overall assessment-level concepts for the presence of ‘child’ OMOP concepts. When ‘child’ concepts were present, raters used Athena to view the OMOP concept hierarchy and search for associated subscale- and item-level concepts. Finally, for complex assessments with publicly accessible details on individual subscales and items, we searched Athena using subscale and item names and/or text.

For DMH data elements, OMOP concepts were considered appropriate to map if they met one of three criteria: (1) they were equivalent to the general phenomenon represented by the data element explicitly or via equivalent terminology, (2) they included the data element phenomenon as a direct subcategory or list component, or (3) they represented the data element phenomenon with an additional specification of diagnostic subtype, data source, time frame, or survey item text eliciting information on the data element phenomenon. As done in previous OMOP mapping studies (e.g., Cai et al., 2023),^25^ raters labeled each mapping using a modified version of the Health Level 7 Fast Healthcare Interoperability Resources (FHIR) concept-map equivalence scale,^26^ with levels reflecting each of the above three criteria (OMOP concept *equivalent* to, *broader* than, or *narrower* than the corresponding data element; **Supplemental Table 1**). Due to the conceptual broadness of many DMH data elements, raters were limited to 20 mappings per data element at the initial mapping stage.

For assessment data elements, OMOP concepts were considered appropriate to map only if they pertained to the same assessment *and* level of granularity (i.e., overall assessment-level vs. subscale-level vs. item-level). OMOP concepts representing the correct assessment, but a different level of granularity (e.g., an OMOP concept representing the overall ARAT vs. the ARAT item-level data element) were not considered appropriate to map.

### 2.3. Initial Mapping Agreement Metrics and Associated Statistical Analyses

Analysis of rater agreement followed the process used by French et al., 2024.^17^ For both data element categories, initial mapping results between raters were analyzed via two metrics: (1) OMOP inclusion agreement and (2) OMOP concept agreement. For OMOP inclusion agreement, we calculated counts and percentages for data elements that were mapped by both raters, only one rater, and neither rater. For OMOP concept agreement, we pooled OMOP concepts for data elements mapped by both raters, and calculated counts and percentages for concepts that were used by both raters and only one rater. For DMH data elements, we also analyzed OMOP concept agreement exclusively for mappings labeled as *equivalent* to determine agreement on a more focused set of mappings. For each agreement metric, we calculated Cohen’s kappa^27^and Gwet’s agreement coefficient (Gwet’s AC),^28,29^ with the latter serving as the primary outcome due to the known limitations of Cohen’s kappa for capturing high rater agreement with heavily unbalanced observations (e.g., a high percentage of positive mapping results).^29-32^ For Gwet’s AC, > 0.8 indicates very good agreement, 0.61 to 0.8 indicates good agreement, 0.41 to 0.6 moderate agreement, 0.21 to 0.4 fair agreement, and ≤ 0.2 poor agreement. All agreement analyses were performed using R (version 4.3.3)^33^ and the dplyr package (version 1.1.4),^34^ with statistical tests completed via the irrCAC package (version 1.4).^35^

### 2.4. Mapping Reconciliation and Final Agreement

All initial mapping differences were discussed between raters to reach full agreement and produce final mapping lists. Specifically, raters reviewed: (1) data elements that were mapped only by one rater and (2) individual OMOP concept mappings completed by only one rater. Athena was consulted as needed during the reconciliation process to review OMOP concept details, including associated hierarchical data structures (e.g., to determine if an OMOP concept was mapped to the correct item-level assessment data element or to determine whether OMOP concepts in the ‘Meas Value’ domain correctly represented data elements vs. representing an answer to an unrelated survey item).

### 2.5. Final Mapping Analysis

The final reconciled DMH and assessment mapping lists were descriptively analyzed to characterize the breadth and depth of OMOP coverage of ENIGMA-SR data elements. We calculated counts and percentages for OMOP inclusion of DMH data elements and distinct assessments. For complex assessments, we calculated counts and percentages for OMOP inclusion of subscale- and item-level data elements, also examining the highest level of data element granularity in OMOP (assessment-, subscale-, or item-level) and comparing it to the highest granularity levels of ENIGMA-SR database variables. Finally, we calculated descriptive statistics (median, range and counts/percentages) for mapped OMOP concepts per data element, as well as the OMOP *domains* in which concepts resided (i.e., the storage location within the data model). All final mapping analyses and data visualizations were performed using R (version 4.3.3),^33^ with dplyr (version 1.1.4),^34^ gtsummary (version 2.1.0),^36^ and ggplot2 (version 3.5.2)^37^ packages.

## 3. Results

### 3.1. Initial OMOP Inclusion Agreement

During the initial mapping process of DMH data elements (n = 46), 76.1% (35) were mapped by both raters, 15.2% (7) were mapped by only one rater, and 8.7% (4) were unmapped by both raters, for a total agreement rate of 84.8% and ‘good’/bordering ‘very good’ overall agreement based on Gwet’s AC (0.79). The disparity between Gwet’s AC and Cohen’s kappa (0.45) here highlights the limitations of Cohen’s Kappa discussed above.

For assessment data elements (n = 200), 34.0% (68) were mapped by both raters, 6.0% (12) were mapped by only one rater, and 60.0% (120) were unmapped by both raters, for a total agreement rate of 94.0% and metrics indicating ‘very good’ overall agreement (Gwet’s AC: 0.89; Cohen’s kappa: 0.87).

### 3.2. Initial OMOP Concept Agreement and Reconciliation

For DMH data elements initially mapped by both raters, 255 total unique OMOP concepts were used across the three equivalency levels (broader, equivalent, narrower). Of these, 37.6% (96) were mapped by both raters and 62.4% (159) were mapped by only one rater, for a ‘poor’ overall agreement between raters (Gwet’s AC: -0.09; Cohen’s kappa: -0.31). Considering *equivalent* DMH mappings only (n = 104), as expected, agreement improved, with 58.7% (61) mapped by both raters and 41.3% (43) mapped by only one rater. Though, agreement still only measured as ‘fair’/bordering ‘moderate’ based on our primary metric (Gwet’s AC: 0.38; Cohen’s kappa: -0.26). Overall, lack of DMH agreement was driven by a combination of the broadness of DMH data elements, the use of exhaustive mapping, and the redundancy of OMOP concepts for this type of data.

For all assessment data elements initially mapped by both raters (n = 68), 476 total unique OMOP concepts were used. Of these, 56.5% (269) were mapped by both raters and 43.5% (207) were mapped by only one rater, for only ‘fair’ agreement between raters (Gwet’s AC: 0.34; Cohen’s kappa: -0.27). However, most disagreement arose from four specific item-level data elements, three for which disagreement was driven by a disproportionately large quantity of redundant OMOP concepts. With these data elements removed (n = 64), among 274 total unique concepts, 86.5% (237) were mapped by both raters and 13.5% (37) were mapped by only one rater, for ‘very good’ agreement between raters (Gwet’s AC: 0.85; Cohen’s kappa: - 0.05).

Despite some challenges with initial mapping rater agreement for specific OMOP concepts, mapping differences for all data elements were reconciled to reach 100% agreement via discussion between raters, consulting Athena as needed. During the reconciliation process for DMH data elements (across all mapping equivalency levels), 24/274 were determined to be incorrect, leaving 250 final mappings (*broader* = 19; *equivalent* = 98; *narrower* = 133). For assessment data elements, 149/527 were determined to be incorrect, leaving 378 final mappings (assessment-level = 120; subscale-level = 18; item level = 240). All final data element mappings are provided in **Supplemental Table 2** and **Supplemental Table 3**.

### 3.3. Overall Inclusion in OMOP

DMH data elements (n = 46) had fairly extensive OMOP coverage, with 84.8% (39) having at least one OMOP concept mapped (at *any* mapping equivalency level). Unmapped DMH data elements spanned 3 data categories (care/study-related information, demographics, stroke-specific medical history) and all were stroke-related (**Table 3**). When considering only mappings labeled as *equivalent*, OMOP coverage fell to 76.1% (35, eliminating 4 data elements with only *narrower* mappings), and most data elements losing coverage (3/4) were, again, stroke-related.

**Table 3.** OMOP inclusion of demographics and medical history (DMH) data elements (n = 46) by data category and stroke-relatedness.

| DMH data characteristics | Data elements | OMOP inclusion by mapping equivalency level |  |
| --- | --- | --- | --- |
|  |  | All levels<sup>1</sup> | Equivalent only<sup>1</sup> |
| Data category |  |  |  |
| Demographics | 16 | 14 (87.5%) | 13 (81.3%) |
| Stroke-related medical history | 12 | 8 (66.7%) | 6 (50.0%) |
| Other medical history | 11 | 11 (100.0%) | 11 (100.0%) |
| Care/study-related information | 7 | 6 (85.7%) | 5 (71.4%) |
| Stroke-related construct |  |  |  |
| Yes | 21 | 14 (66.7%) | 11 (52.4%) |
| No | 25 | 25 (100.0%) | 24 (96.0%) |

<sup>1</sup>n (%)

Distinct assessments (n = 95) had much lower coverage, with only 58.9% (56) having at least one OMOP concept mapped (at *any* data element granularity level). Psychosocial assessments had the most coverage (69.2%) and cognitive assessments the least (51.5%; **Table 4**). Complex assessments (n = 73), or assessments that had multiple items and/or generated multiple datapoints, had an almost 20 percentage point higher OMOP inclusion rate than non-complex assessments (n = 22; 63.0% vs. 45.5%).

**Table 4.** OMOP inclusion of rehabilitation assessments by assessment category, stroke-specificity, and complexity.

| Assessment characteristics | Assessments (N = 95) | OMOP inclusion<sup>1</sup> |
| --- | --- | --- |
| Assessment category |  |  |
| Cognitive | 33 | 17 (51.5%) |
| Global function | 16 | 10 (62.5%) |
| Psychosocial | 13 | 9 (69.2%) |
| Sensorimotor | 33 | 20 (60.6%) |
| Stroke-specific assessment |  |  |
| No | 90 | 54 (60.0%) |
| Yes | 5 | 2 (40.0%) |
| Complex assessment |  |  |
| Yes | 73 | 46 (63.0%) |
| No | 22 | 10 (45.5%) |

<sup>1</sup>n (%)

### 3.2. Depth of OMOP Coverage for Complex Assessments

For complex assessments (n = 73), we explored the depth of OMOP coverage for subscale-, and item-level data elements, comparing it to that of ENIGMA-SR variables. Subscale- and item-level data elements had far lower OMOP inclusion rates than assessment-level data elements (**Table 5a**). Additionally, these inclusion rates were lower than those of ENIGMA-SR variables, though when only considering complex assessments included in OMOP (at *any* granularity level; n = 46), differences between OMOP and ENIGMA-SR item-level inclusion rates evaporated (both at 30.4%; **Table 5b**). Subscale-level data elements had particularly poor OMOP coverage, which was consistently lower than ENIGMA-SR, regardless of which group of complex assessments was considered (**Table 5a,b**).

**Table 5.**
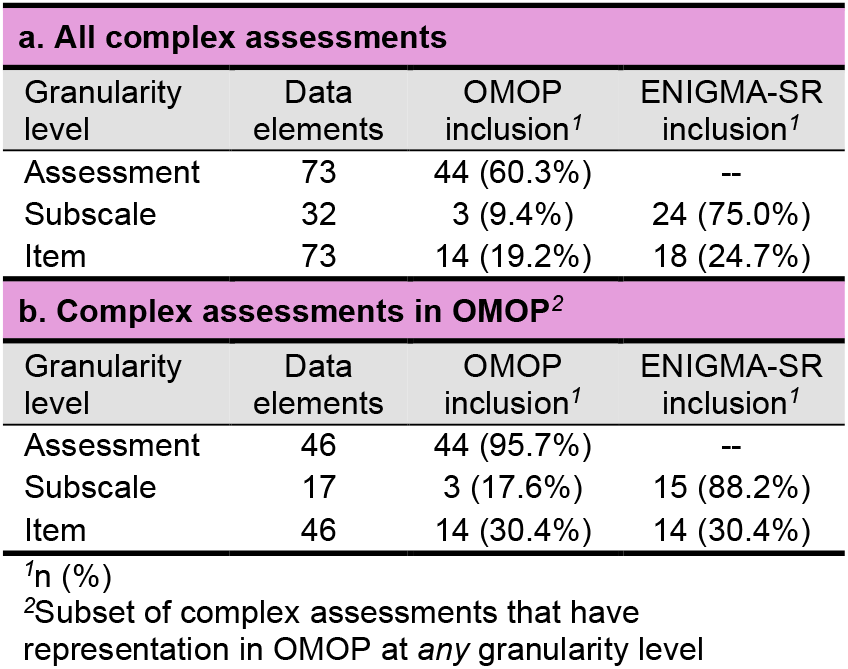
OMOP vs. ENIGMA-SR database inclusion of complex assessment granularity levels.

| a. All complex assessments |  |  |  |
| --- | --- | --- | --- |
| Granularity level | Data elements | OMOP inclusion<sup>1</sup> | ENIGMA-SR inclusion<sup>1</sup> |
| Assessment | 73 | 44 (60.3%) | -- |
| Subscale | 32 | 3 (9.4%) | 24 (75.0%) |
| Item | 73 | 14 (19.2%) | 18 (24.7%) |
| b. Complex assessments in OMOP<sup>2</sup> |  |  |  |
| Granularity level | Data elements | OMOP inclusion<sup>1</sup> | ENIGMA-SR inclusion<sup>1</sup> |
| Assessment | 46 | 44 (95.7%) | -- |
| Subscale | 17 | 3 (17.6%) | 15 (88.2%) |
| Item | 46 | 14 (30.4%) | 14 (30.4%) |

<sup>1</sup>n (%)
<sup>2</sup>Subset of complex assessments that have representation in OMOP at *any* granularity level

Next, we determined what proportion of complex assessments OMOP covered at sufficient depth to match ENIGMA-SR. Specifically, for each assessment, we compared the highest granularity level represented in OMOP to that of data included in ENIGMA-SR (granularity levels, lowest to highest: overall assessment-level, subscale-level, item-level). OMOP met or exceeded ENIGMA-SR granularity for 43.8% (n = 32) of assessments, falling short for the remaining 56.2% (n = 41). Even when only considering complex assessments included in OMOP (n = 46), OMOP still fell short for 30.4% (n = 14) of assessments.

### 3.5. Mapped OMOP concept averages and characteristics

For DMH mappings (data elements: 39; OMOP concepts: 250) the median number of OMOP concepts per data element was 6 (range: 1-23). Medians and ranges for OMOP concepts mapped per data element generally increased across mapping equivalency levels from *broader* to *narrower*, as expected for a hierarchical, taxonomy-like structure (**Figure 1**). OMOP concepts most frequently resided in the OMOP domains of Observation at 41.6% (104), Condition at 36.8% (92), and Meas Value at 15.2% (38). Importantly, among DMH data elements mapped to multiple OMOP concepts (n = 31), 58% (18) had concepts within 2 to 3 different OMOP domains (**Figure 2a**). This was true even when only considering mappings labeled as *equivalent*: 60.9% (14/23) of multi-mapped data elements. See **Supplemental Figure 1** for individual DMH data element concept and domain counts.

**Figure 1.**
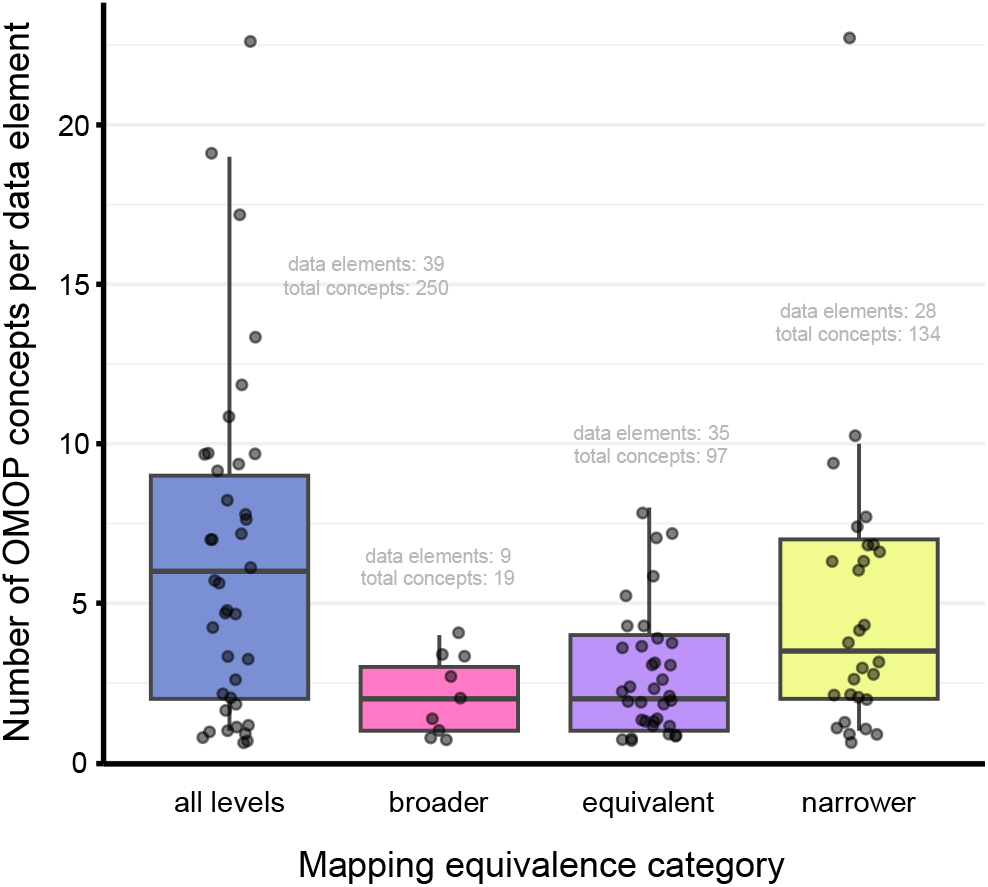
Number of OMOP concepts per mapped demographics and medical history (DMH) data element by mapping equivalency category. Note: mapping equivalency categories are assigned per individual mapping (*not* per data element). As each data element can be mapped to multiple OMOP concepts, it can be associated with multiple mapping equivalence categories.

**Figure 2.**
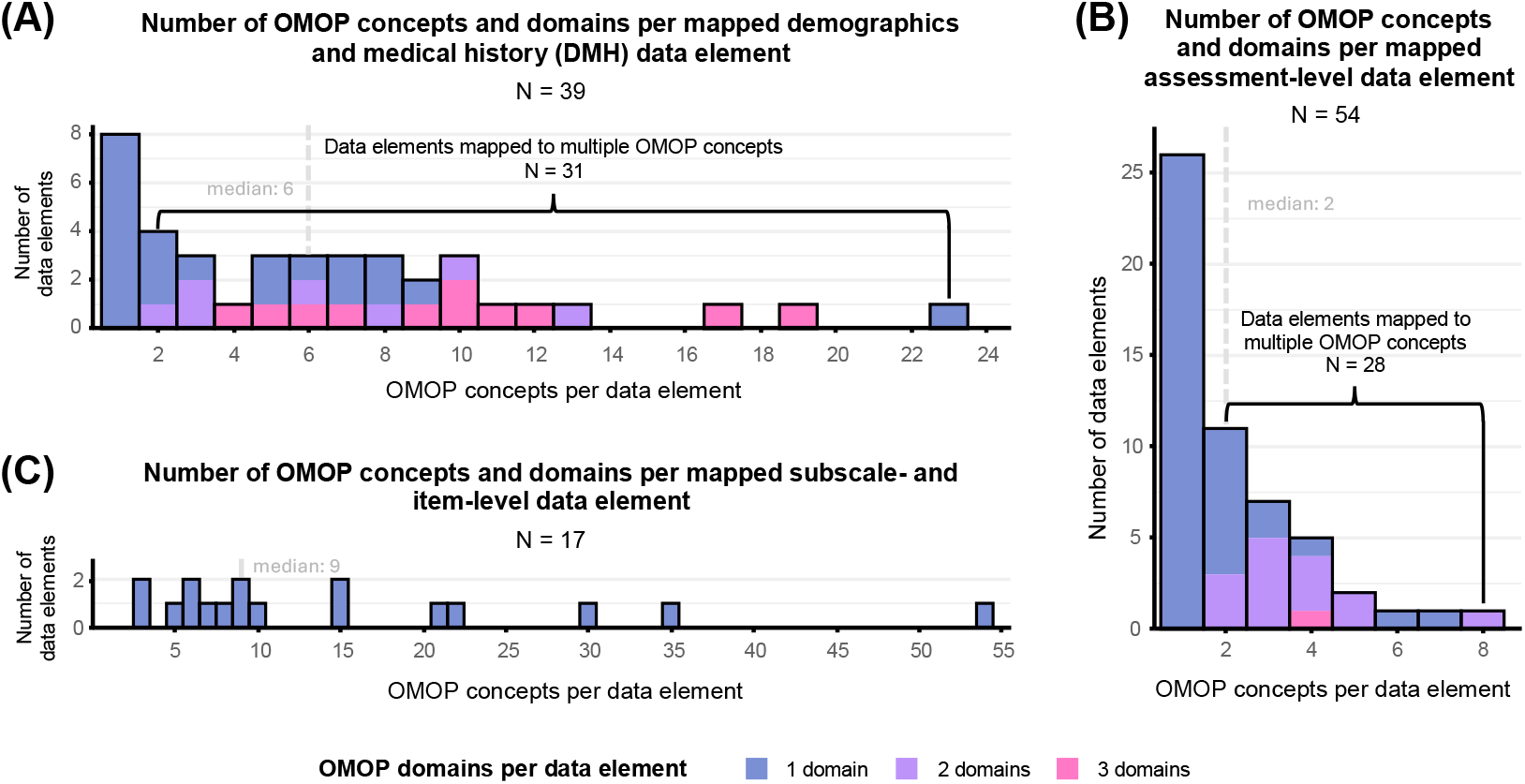
Number of OMOP concepts and domains per mapped (A) demographics and medical history (DMH) data element, (B) assessment-level data element, and (C) subscale- and item-level assessment data elements. Note: each subscale- and item-level data element represents *all* subscales and items comprising a single assessment

For the assessment category, overall assessment-level mappings (data elements: 54; OMOP concepts: 120) had a median of 2 (range: 1-8) concepts per data element. OMOP concepts mapped to these data elements most frequently resided in the OMOP domains of Measurement at 66.7% (80) and Observation at 25.8% (31). Similar to DMH data elements, among assessment-level data elements mapped to multiple OMOP concepts (n = 28), 53.6% (15) had concepts within 2 or more different OMOP domains (**Figure 2b**). See also **Supplemental Figure 2** for individual assessment-level data element concept and domain counts.

For subscale- and item-level mappings (data elements: 17; OMOP concepts: 258) the median number of OMOP concepts per data element increased to 6 (range: 3-9) and 12.5 (range: 3-54), respectively, reflecting the hierarchical structure of complex assessments. Contrary to assessment-level mappings, OMOP concepts for subscale- and item-level mappings resided most frequently in the Observation domain at 81.8% (211) followed by Measurement at 18.2% (47), and all concepts for each data element were all within a single OMOP domain (**Figure 2c**). All subscale-level OMOP concepts (n = 18) represented unique assessment subscales (no subscales repeated), while 25% (60) of item-level OMOP concepts (n = 240) represented duplicate assessment items. These duplicates most often had different specifications for data produced by the same item, such as assessment timeframe (IRF-PAI Mobility, IRF-PAI Self-Care) or variations in how item responses can be recorded (Patient Health Questionnaire 9).

## 4. Discussion

In this study, we characterized the breadth and depth of OMOP CDM coverage for two general categories of data within a large-scale stroke rehabilitation research database: (1) demographics and medical history (DMH) and (2) rehabilitation-related assessments. Consistent with other recent rehabilitation data mapping efforts,^17^ initial mapping agreement between raters was good to very good for DMH and assessment data element OMOP inclusion, as well as for specific OMOP concepts for overall assessment- and subscale-level assessment data elements. Initial agreement was variable for specific OMOP concepts for DMH and item-level assessment data elements; however, raters were able to reconcile to full agreement to produce a final mapping list.

We found that DMH data elements generally had more inclusion in OMOP than did assessments (84.8% vs. 58.9% overall). Though assessment inclusion was still noticeably higher than for the neurologic and orthopedic rehabilitation assessments mapped by French et al. in 2024 (46.9% and 48.1%),^17^ which may be due to assessment pool differences and/or updates to OMOP vocabularies between studies. For DMH data elements, being stroke-related was the primary factor associated with OMOP exclusion, while for assessments, cognitive assessments were the most excluded and psychosocial assessments the most represented. For complex assessments (including multiple items and/or datapoints), OMOP had very limited coverage of subscale- and item-level data elements (9.4% and 19.2%), and the granularity of OMOP representation for these assessments did not match that of ENIGMA-SR database for 56.2% of assessments (30.4% when only considering assessments included in OMOP). Furthermore, several key assessments central to stroke recovery research have poor coverage or are completely missing in OMOP. For example, the Fugl-Meyer Upper Extremity Motor Assessment (FM-UE), Wolf Motor Function Test (WMFT), and ARAT are three of the most frequently reported measures in ENIGMA-SR and are among the upper extremity outcome measures demonstrating the strongest measurement quality and utility for stroke recovery research.^38,39^ Yet, the FM-UE and WMFT have no OMOP coverage at all and the ARAT has only assessment-level coverage.

Critically, similar to the findings of French et. al, 2024,^17^ we observed considerable variability in data element representation within the OMOP CDM structure. DMH and overall assessment-level assessment data elements were commonly mapped to multiple OMOP concepts, and over half of such data elements had concepts residing in multiple different OMOP domains. Additionally, they showed variability predominantly in *where* they were represented within the CDM, as the domains were almost always Measurement and Observation (indicating data fields within distinct data tables). For DMH data elements, variability was more complex, as a higher proportion were mapped to 3 domains (most often Observation, Condition, and Meas Value), and these different domains produced variability in both *where* and *how* data elements were represented in the CDM. Condition and Observation both designate specific data tables, while Meas Value concepts can exist in multiple tables (*where*). The Observation domain designates a field to which values are linked within a table, while Condition and Meas Value domains assert the value itself (*how*). For example, the data element ‘Myocardial Infarction (MI)’ could be mapped to one of three OMOP concepts sharing the same exact name (‘Myocardial infarction’; **Supplemental Table 1**): OMOP concept 40761359 (Observation domain) represents a data *field* within the Observation table, concept 4329847 (Condition domain) represents a data *value* (assertion) within the Condition table, and concept 45881971 (Meas Value domain) represents a data *value* (e.g., survey response) within either the Observation table or Measurement table. This variability presents significant roadblocks to data harmonization as it increases the likelihood that the CDM will be applied differently across datasets.

### 4.1. Considerations for using the OMOP CDM for research data specifically

Additionally, the results of this study highlighted several key areas of consideration for the future use of the OMOP CDM specifically to facilitate syntheses of research- and clinical-generated rehabilitation data: (1) data representation, (2) data structure, and (3) limitations of available tool.

#### Data representation

As OMOP was developed to handle clinical data, there are important differences in how it represents data compared to a research database like ENIGMA-SR, which create challenges beyond the general lack of rehabilitation-related concepts. First, there is inherently less direct coverage of research study-related data (e.g., ‘Treatment Group’) and condition specific data (e.g., ‘Stroke Chronicity’, ‘Total Number of Strokes’, ‘Handedness – Post-stroke’). Second, data granularity is often inherently imbalanced between research and clinical data. For example, as our findings demonstrate, the limited coverage of subscale- and item-level assessment data in OMOP contrasts with data collected for studies answering specific research questions that are more accurately evaluated at granular data levels. This disparity could worsen in the future as high-capacity data storage becomes increasingly accessible, motivating researchers to universally store more granular data. Conversely, medical history data in research databases is often less granular than EHR data, as it is often a control or contextual variable based on self-report, rather than a precise test result or diagnosis. This is especially true for pre-harmonized databases such as ENIGMA-SR, where there is already some loss of precision from combining data collected by studies with differing approaches. In our mapping results, this manifested as 4 DMH data elements only being mapped to *narrower* OMOP concepts, which related to specific survey questions or sources (affected data elements: ‘Date of Stroke’, ‘Smoking – Cigarettes per Day’), or specified an anatomical site or condition subtype (affected data elements: ‘Motor Paresis’, ‘Tissue Plasminogen Activator [tPA]’). Similarly, we found that in OMOP some complex assessment item-level concepts and simple assessment concepts had additional specifications based on a clinical/reporting standard or EHR form that is not relevant to research data from the same assessment (e.g., the specified timeframe for an IRF-PAI Self-Care item, ‘Upper body dressing - functional ability during 3 day assessment period [CMS Assessment]’ [ID: 36204530]; the specific anatomical location for ‘Monofilament foot sensation test’ [ID: 4047085]). Ultimately, such granularity mismatches will need to be carefully considered when harmonizing clinical and research data, as data will have to be represented at the least specific level between the two.

#### Data structure

Similarly, there are significant structural differences between clinical data typically handled by OMOP and data housed within a research databases such as ENIGMA-SR. Primarily, research data is usually stored in a ‘flat’ format (e.g., a single CSV file) for ease of use and compatibility with statistical software, while EHR data is typically hierarchical or nested (e.g., flowsheets, linked tables), often using proprietary data forms fulfilling specific clinical or reporting needs. This contrast in structural complexity presents several challenges to accurate and efficient mapping, the first of which is ambiguity in selecting the most appropriate OMOP concept. As discussed in our results, the complexity of the OMOP CDM is accompanied by a high degree of flexibility in how rehabilitation data can be represented (likely related to variations in EHR data structures). When attempting to merge this level of complexity and variability with the relative simplicity of research data, specific mapping decisions can seem borderline arbitrary. Next, OMOP represents data hierarchies via formal vocabulary structures, while research databases often indirectly represent them via variable names and external descriptions. For example, for the Wechsler Adult Intelligence Scale, OMOP uses linked hierarchical concepts: ‘Wechsler Adult Intelligence Scale 4th Edition (WAIS-IV)’ (ID: 40219612; assessment-level), ‘WAIS-IV: Perceptual Reasoning Index (PRI)’ (ID: 40219497; subscale-level), and ‘WAIS-IV: Perceptual Reasoning Index (PRI) - Matrix Reasoning’ (ID: 40219500; item-level), where each of the latter two is a child of the previous. Conversely, a research database such as ENIGMA-SR may use the variable names: WAIS, WAIS_PRI, and WAIS_PRI_MATRIX, supplemented by a data dictionary. These implied hierarchical structures are easily obscured when research data does not populate every level of the hierarchy, as occurs with assessments not having a singular total/composite score, or when studies only collect certain assessment items or subscales. This is apparent in ENIGMA-SR for assessments such as the Comprehensive Aphasia Test Battery, for which the database only includes subscale- and item-level variables. In fact, ENIGMA-SR does not include assessment-level variables for 18 (of 73) complex assessments. Comparatively, 12 of these 18 assessments (66.7%) have assessment-level concepts in OMOP. A final related consideration is differences in variable and concept naming conventions, especially for assessment item-level data. For example, OMOP often directly uses assessment item text as concept names (versus the abbreviated labels and acronyms often used for research variables). Additionally, some of these item-level names do not explicitly include the name of the parent assessment (e.g., ‘Lack of energy’ [ID: 1761073] is a child concept under ‘Center for Epidemiologic Studies Depression Scale panel [CES-D]’ [ID: 1761569]), which increases the difficulty of locating these concepts while mapping research data.

#### Limitations of available tools

Widely used open-source tools intended to ease the arduous process of OMOP mapping (i.e., USAGI and Athena) are limited in their ability to address the above research data-specific challenges. First, no current tool provides a timely solution to the lack of coverage of critical research outcomes (whether they are completely missing or have only minimal representation), more niche condition-specific data, or study-related data. Second, USAGI’s term similarity approach does not effectively overcome challenges arising from the combined differences in data structure and naming conventions between OMOP and research databases like ENIGMA-SR. Specifically, as noted in previous studies, while USAGI’s automated recommendations for more concrete, finite medical data (e.g., medications) tend to be accurate, its algorithm struggles to provide accurate recommendations for phenomena with more variability in terminology and specificity (e.g., conditions, other more broader medical terms).^40-42^ Throughout our own mapping process, automated recommendations were often incorrect, and frequently concepts more closely related to a given data element had lower ‘match scores’ than concepts only loosely related. This was particularly true for assessments or assessment items having long, complex names or multiple versions/iterations (common in rehabilitation data). Ultimately, accurate concept mapping for our proxy data elements required a substantial amount of manual concept exploration and mapping, and it is highly probable that an even more substantial effort would have been required if using the original compact research-based variable names. USAGI’s limitations are tied to its use of a lexical approach based purely on literal text similarity (TF-IDF),^23^ leaving it virtually unable to evaluated semantic similarity (i.e., different words with the same meaning). It is clear that more robust tools are needed to handle the complexity of typical rehabilitation-related data, especially when it originates from a research-based source. Fortunately, researchers have begun to implement and evaluate alternate approaches based on emerging AI technologies such as deep learning and Large Language Models (LLMs), with varying levels of success.^40-45^

### 4.2. Future directions

Building on these findings, several avenues can be explored to continue to work toward the ultimate goal of large-scale synthesis of clinical- and research-generated rehabilitation data. As others have suggested,^12,16,17^ advocacy for the expansion of OMOP to include more rehabilitation-related assessments and data will be critical to harmonizing large-scale rehabilitation datasets (from any source) with meaningful levels of granularity. The unmapped concepts identified in this study will be shared with the Observational Health Data Sciences and Informatics (OHDSI) Rehabilitation Workgroup to support ongoing community-driven efforts to expand OMOP vocabulary coverage of rehabilitation-related constructs. Moreover, building conceptual maps and cross-walks to research-focused data standards, such as the NINDS NeuroRehab CDEs,^11^ can further aid the future synthesis of this type of data with EHR-produced data. In addition, exploring the feasibility of adapting emerging deep learning and LLM-based tools to rehabilitation-related data (and even more specifically for research-generated rehabilitation data), may significantly ease the current burden of arduous mapping processes, especially if resulting maps are used as the gold standard for model training. Finally, if this proves successful, it may be worth exploring the integration of these adapted tools with other rehabilitation-specific EHR data extraction tools currently being developed.^12^

### 4.3. Limitations

While our findings greatly advance knowledge on the practicality of using a clinical-focused CDM such as OMOP to facilitate harmonization of rehabilitation research data, there are several limitations. First, ENIGMA-SR is a condition-specific database that has evolved in purpose, content, and scale over the years, and whose primary metric is neuroimaging. Therefore, the data within ENIGMA-SR is not necessarily representative of data that might be in other large-scale rehabilitation research databases, especially those focused on dissimilar conditions. Additionally, we did not use a one-to-one representation of ENIGMA-SR assessment data, which, while enabling a more structured analysis and interpretable results, does not account for the coverage of each individual subscale and item comprising complex assessments (for OMOP or ENIGMA-SR). Relatedly, representations of and mapping processes for subscale- and item-level assessment data elements were based on publicly available information (i.e., databases, manuals, publications), which may have limited the precision of our analyses for assessments with limited publicly available information. We also did not explicitly account for the frequency at which assessments have been captured in ENIGMA-SR, instead giving each assessment equal weight in our analyses, regardless of whether 1 or 60+ ENIGMA-SR studies collected it. Next, as we did not account for anatomical location/laterality or variations in measurement units/assessment scoring via the proxy data elements used for this study, we are unable to evaluate OMOP coverage related to these phenomena, as others have done (e.g., Cai, et al., 2023).^25^

## 5. Conclusion

We found that the OMOP CDM has strong coverage of general demographics and medical history-related data, with far less coverage of stroke-specific data and rehabilitation-related assessments, especially at more granular data levels. Furthermore, for the coverage that exists, there is a high degree of variability in OMOP concept representation and designated CDM location, especially for DMH and overall assessment-level data. Through our analyses, we identified key considerations for the future use of the OMOP CDM to synthesize clinical and research rehabilitation data related to data representation and structure, as well as the limitations of current tools supporting mapping processes. Moving forward, it will be critical to improve the breadth and depth of OMOP coverage of rehabilitation-related data. Additionally, large-scale research-clinical rehabilitation data synthesis may be greatly aided by establishing connections between OMOP and research-based data guidelines and exploring the capability of emerging technologies to help develop innovative support tools.

## Data Availability

All data produced in the present work are contained in the manuscript

