## Supplements for "Exploring the Application of the Observational Medical Outcomes Partnership Common Data Model to Multi-site Stroke Rehabilitation Research Data"

### SUPPLEMENTAL DOCUMENTS

**Supplemental Document 1.** Initial OMOP Mapping Procedures

#### Data preparation

- ENIGMA-SR variables have been collapsed into 95 assessments and 46 demographics and medical history data elements
- Variables divided into two separate CSVs
  - Demographics and medical history (n = 46)
    - One row per data element
  - Assessments (n=200)
    - Includes data elements for top-level assessments, as well as for assessment subscales and items for those that have subscales and/or multiple items
    - Each level (overall assessment, subscale, and item) is represented by a single data element

#### Import data into USAGI (for each data category CSV)

- Column mapping:
  - Source code column = N/A
  - Source name column = Data OR Assessment
  - Additional info column = N/A
- Filters:
  - Standard concepts
  - Include source terms

#### Map OMOP standard concept IDs with USAGI (for data element within each CSV)

- Look at the suggested concept ID to make sure it is appropriate to map to the data element
- Review the list of other potential concept ID matches and map all others that are also appropriate
- For demographics and medical history variables
  - Up to **20** concept IDs can be mapped per data element
  - If there are more than 20 concept IDs that apply to the data element, choose the top 20 that you feel are most relevant
- For assessment variables
  - For each assessment level (overall assessment, subscales, items), map **all** appropriate CIDs (e.g., all overall assessment CIDs to the assessment variable, all subscale CIDs to the assessment subscale-level variable, all item CIDs to the assessment item-level variable)
  - For subscale- and item-level data elements:
    - Look through the list of recommended CIDs for any that match (do not assume there are not subscale-level or item-level CIDs just because there is no overall assessment CID)

        - Look at the overall assessment CID(s) for ‘children’. If the CID has any, look up the CID in the Athena database (https://athena.ohdsi.org/search-terms/terms) by CID number. If there are multiple ‘children’, they are likely CIDs for subscales or items of that assessment. After locating them in Athena, copy/paste each full name into the USAGI ‘Query’ field to map them. This step is important due to the limitations of the USAGI platform in identifying subscale- and item-level CID matches.

- Appropriateness for mapping
    - The likeness score that USAGI provides can be used as a reference, but should not be used as the primary decider of whether a concept ID should be mapped to a variable
    - **For demographic and medical history data elements:** the concept should represent the same general phenomenon as the data element, an umbrella phenomenon directly related to or including that phenomenon (e.g., Dyslipidemia CID for High Cholesterol data element), or a very broad subtype of that phenomenon (e.g., Diabetes Type 2 CID for the Diabetes data element, or Left Hemiplegia CID for the Motor Paresis data element). Do NOT map very specific subtypes of the data element phenomenon (e.g., specific sequelae of a condition or very specific organs/anatomical regions related to a condition)
    - **For assessment data elements:** the CID should pertain to the same level of granularity as the variable (i.e., total assessments/total assessment scores are only mapped to concepts representing the total assessment, sub-scores are only mapped to corresponding sub-score concepts, items are only mapped to corresponding item concepts)

**<u>Export mapping results from USAGI (for each CSV)</u>**

- Use the ‘File’ > ‘Export for review’ option
- Export all (Approved and Unapproved) mappings

**<u>Categorization of demographics and medical history mappings based on equivalency</u>**

- After exporting the mapping results for demographics and medical history, add an additional column to the CSV labeled ‘map_cat’. For each mapping, add labels “broader”, “equivalent”, or “narrower”
    - **broader** – the CID is an umbrella category directly related to the data element phenomenon (e.g., Dyslipidemia CID for High Cholesterol data element) or a list that includes the data element phenomenon (e.g., Race or Ethnicity CID for the Race data element).
    - **equivalent** – the CID directly represents the data element explicitly or via an equivalent regional term (e.g., Dyspraxia CID for the Apraxia data element)
    - **narrower** – the CID represents the data element phenomenon with additional specification of diagnostic subtypes (e.g., Diabetes Type 2 CID for the Diabetes data element), data source (e.g., Ethnicity - calculated or BMI [estimated] CIDs), or timeframe (e.g., ‘Myocardial infarction in past 24 hours’ CID for Myocardial Infarction data element), or the CID is a specific survey item eliciting the data element phenomenon (e.g., “How many cigarettes per day do, or did, you smoke” CID)
- Leave unmapped rows blank for ‘map_cat’

### SUPPLEMENTAL TABLES

**Supplemental Table 1.** Equivalency categories for mapping demographics and medical history (DMH) data elements to Observational Medical Outcomes Partnership (OMOP) standard concepts

| Equivalency category[1] | Qualifier | Example mapped OMOP concept | Example mapped data element |
|---|---|---|---|
| **broader** | OMOP concept includes the data element as a direct subcategory | ‘Dyslipidemia’ (ID: 4159131) | ‘High Cholesterol’ |
| | OMOP concept includes the data element explicitly in a list | ‘Race or ethnicity’ (ID: 3050381) | ‘Ethnicity’ |
| **equivalent** | OMOP concept explicitly represents data element | 'Age’ (ID: 3022304) | ‘Age’ |
| | OMOP concept represents a phenomenon equivalent to data element | ‘Dyspraxia’ (ID: 4290260) | ‘Apraxia’ |
| **narrower** | OMOP concept has additional specification of diagnostic subtype | 'Diabetes Type 2’ (ID: 45877606) | ‘Diabetes’ |
| | | ‘Left hemiparesis’ (ID: 4081504) | ‘Motor Paresis’ |
| | OMOP concept has additional specification of data source or standard | ‘Tabulated ethnicity [CDC]’ (ID: 21494233) | ‘Ethnicity’ |
| | | ‘ECG: atrial fibrillation’ (ID: 4064452) | ‘Atrial Fibrillation (AFib)’ |
| | OMOP concept has additional specification of a timeframe | 'Myocardial infarction in past 24 hours’ (ID: 21491677) | ‘Myocardial Infarction (MI)’ |
| | OMOP concept is a specific survey item eliciting the data element phenomenon | ‘Ever told by doctor or nurse that you have high blood cholesterol’ (ID: 40761393) | ‘High Cholesterol’ |

[1]Categories based on the Health Level 7 Fast Healthcare Interoperability Resources (FHIR) concept-map equivalence scale

**Supplemental Table 2.** Final mapping of demographics and medical history (DMH) data elements to Observational Medical Outcomes Partnership (OMOP) standard concepts

| **A. Demographics** | | | | | |
|---|---|---|---|---|---|
| **Data Element** | **Mapping equivalency level** | **OMOP Concept Name** | **OMOP Concept ID** | **OMOP Domain** | **Source Vocabulary** |
| Age | Equivalent | Age | 4265453 | Meas Value | SNOMED |
| | | Age | 3022304 | Observation | LOINC |
| | | Current chronological age | 4314456 | Observation | SNOMED |
| | Narrower | Age - Reported | 3007191 | Observation | LOINC |
| | | Age calculated | 3023631 | Observation | LOINC |
| | | Age estimated | 3008232 | Observation | LOINC |
| Alcohol Consumption - Drinks per Week | Equivalent | Alcohol units consumed per week | 35609491 | Observation | SNOMED |
| | Narrower | Pattern of alcohol consumption through week | 4027353 | Observation | SNOMED |
| | | Pattern of alcohol consumption through week - finding | 4275494 | Observation | SNOMED |
| Body Mass Index | Equivalent | Body mass index | 4245997 | Measurement | SNOMED |
| | | Body mass index (BMI) [Ratio] | 3038553 | Measurement | LOINC |
| | Narrower | Body mass index (BMI) [Percentile] | 40762636 | Measurement | LOINC |
| | | Body mass index (BMI) [Ratio] Estimated | 36304833 | Measurement | LOINC |
| | | Body mass index prime ratio | 37160500 | Measurement | SNOMED |
| Date of Birth | Equivalent | Birth date | 3022007 | Observation | LOINC |
| | | Date of birth | 4083587 | Observation | SNOMED |
| Ethnicity | Broader | Race or ethnicity | 3050381 | Observation | LOINC |
| | | Race or ethnicity OMB.1997 | 40762453 | Observation | LOINC |
| | | Race or ethnicity [The Position Generator] | 40769917 | Observation | LOINC |
| | Equivalent | Ethnic group | 4271761 | Observation | SNOMED |
| | | Ethnicity | 44803968 | Observation | SNOMED |
| | | Ethnicity / related nationality data | 4087925 | Observation | SNOMED |
| | Narrower | Derivation method Computed Ethnicity | 3027483 | Measurement | LOINC |
| | | Ethnicity Computed | 3026646 | Observation | LOINC |
| | | Ethnicity OMB.1997 | 40771985 | Observation | LOINC |
| | | Ethnicity [AHRQ] | 44816592 | Observation | LOINC |
| | | Ethnicity [CDC] | 21494074 | Observation | LOINC |
| | | Ethnicity [USSG-FHT] | 3051581 | Observation | LOINC |
| | | Tabulated ethnicity [CDC] | 21494233 | Observation | LOINC |
| Gender | Broader | Recorded sex or gender | 1988369 | Observation | LOINC |
| | Equivalent | Gender | 4135376 | Observation | SNOMED |
| | | Gender expression | 1450769 | Observation | SNOMED |
| | | Gender expression finding | 1448382 | Observation | SNOMED |
| | | Gender identity | 37171290 | Observation | SNOMED |
| | | Gender identity | 46235215 | Observation | LOINC |
| | | Gender identity finding | 4110772 | Observation | SNOMED |
| | | Gender: Gender Identity | 1585838 | Observation | PPI |
| Handedness - General | Equivalent | Dominant side | 4118850 | Observation | SNOMED |
| | | Handedness | 4211853 | Observation | SNOMED |
| | | Handedness finding | 4270977 | Observation | SNOMED |
| | | Sidedness | 4174140 | Observation | SNOMED |
| | Narrower | Dominant hand [PhenX] | 40768726 | Observation | LOINC |
| Handedness - Post-stroke | N/A | Unmapped | | | |

| A. Demographics | | | | | |
|---|---|---|---|---|---|
| **Data Element** | **Mapping equivalency level** | **OMOP Concept Name** | **OMOP Concept ID** | **OMOP Domain** | **Source Vocabulary** |
| Handedness - Pre-stroke | N/A | Unmapped | | | |
| Race | Broader | Race or ethnicity | 3050381 | Observation | LOINC |
| | | Race or ethnicity OMB.1997 | 40762453 | Observation | LOINC |
| | | Race or ethnicity [The Position Generator] | 40769917 | Observation | LOINC |
| | Equivalent | Race | 4013886 | Observation | SNOMED |
| | | Race | 3046853 | Observation | LOINC |
| | Narrower | Race OMB.1997 | 43055686 | Observation | LOINC |
| | | Race [AHRQ] | 44816591 | Observation | LOINC |
| | | Race [PhenX] | 40759212 | Observation | LOINC |
| | | Race during assessment period [CMS Assessment] | 1259676 | Observation | LOINC |
| Sex | Equivalent | Biological sex | 37116947 | Observation | SNOMED |
| | | Sex | 3046965 | Observation | LOINC |
| | Narrower | Recorded sex or gender | 1988369 | Observation | LOINC |
| | | Sex [AHRQ] | 44816596 | Observation | LOINC |
| | | Sex [USSG-FHT] | 3051551 | Observation | LOINC |
| | | Sex assigned at birth | 46235213 | Observation | LOINC |
| Smoking - Cigarettes per Day | Narrower | Approximately how many cigarettes did you smoke on average [PhenX] | 40770352 | Observation | LOINC |
| | | Cigarettes smoked current (pack per day) - Reported | 3004518 | Observation | LOINC |
| | | How many cigarettes do you smoke per day now [PhenX] | 40766929 | Observation | LOINC |
| | | How many cigarettes per day do, or did, you smoke | 40766364 | Observation | LOINC |
| | | On the average of the entire time you smoked, how many cigarettes did you smoke per day [PhenX] | 40766930 | Observation | LOINC |
| | | On the average, when you smoke, or on the last day you smoked every day, about how many cigarettes do, or did, you smoke [PhenX] | 40766579 | Observation | LOINC |
| | | On the number of days you reported you smoked cigarettes during the past 30 days, how many cigarettes did you smoke per day, on average [PhenX] | 40766353 | Observation | LOINC |
| Smoking - Pack-years | Equivalent | Cigarettes pack-years smoked during life | 903650 | Measurement | OMOP Extension |
| | | Pack years | 4151768 | Measurement | SNOMED |
| | Narrower | Cigarettes smoked total (pack per year) - Reported | 3003421 | Observation | LOINC |
| Smoking Status *current/past* | Equivalent | Lifestyle: Smoking | 1585856 | Observation | PPI |
| | | Smoking | 45880274 | Meas Value | LOINC |
| | | Smoking status | 35811013 | Observation | UK Biobank |
| | | Tobacco smoking | 35811623 | Observation | UK Biobank |
| | | Tobacco smoking status | 43054909 | Observation | LOINC |
| | | Tobacco smoking status for tobacco smoker | 21494888 | Observation | LOINC |
| | Narrower | Have you ever smoked [FACIT] | 42868954 | Observation | LOINC |
| | | Have you ever smoked cigarettes [PhenX] | 40766927 | Observation | LOINC |

| A. Demographics | | | | | |
|---|---|---|---|---|---|
| **Data Element** | **Mapping equivalency level** | **OMOP Concept Name** | **OMOP Concept ID** | **OMOP Domain** | **Source Vocabulary** |
| Smoking Status *current/past (cont.)* | Narrower *(cont.)* | Have you ever smoked part or all of a cigarette [PhenX] | 40766305 | Observation | LOINC |
| | | Smoking status [FTND] | 40766362 | Observation | LOINC |
| Waist-Hip Ratio | Equivalent | Waist/hip ratio | 4087501 | Measurement | SNOMED |
| Years of Education | Equivalent | Schooling | 4144287 | Observation | SNOMED |
| | Narrower | Years of education [#] - Reported | 42528764 | Observation | LOINC |

| B. Stroke-specific Medical History | | | | | |
|---|---|---|---|---|---|
| **Data Element** | **Mapping equivalency level** | **OMOP Concept Name** | **OMOP Concept ID** | **OMOP Domain** | **Source Vocabulary** |
| Affected Side | Equivalent | Affected area | 37117125 | Spec Anatomic Site | SNOMED |
| | Narrower | Side affected [PhenX] | 40768415 | Observation | LOINC |
| Aphasia *presence/history* | Broader | Aphasia, agnosia, dyslexia AND/OR apraxia | 4024716 | Condition | SNOMED |
| | Equivalent | Aphasia | 440424 | Condition | SNOMED |
| | | Aphasia | 45883046 | Meas Value | LOINC |
| | | Dysphasia | 36307452 | Meas Value | LOINC |
| | | Dysphasia | 441594 | Condition | SNOMED |
| | Narrower | Aphasia [Minimum Data Set] | 3043523 | Observation | LOINC |
| | | Aphasia due to brain damage | 46272980 | Condition | SNOMED |
| | | Aphasia in last 7 days [MDSv3] | 40757936 | Observation | LOINC |
| | | Complete aphasia | 4263333 | Condition | SNOMED |
| | | Global aphasia | 4044923 | Condition | SNOMED |
| | | Paraphasia | 4203167 | Condition | SNOMED |
| Apraxia *presence/history* | Equivalent | Apraxia | 132342 | Condition | SNOMED |
| | | Classic apraxia | 4041690 | Condition | SNOMED |
| | | Dyspraxia | 4290260 | Condition | SNOMED |
| | | Motor apraxia | 4202653 | Condition | SNOMED |
| | Narrower | Apraxia as sequela of cerebrovascular accident | 42535344 | Condition | SNOMED |
| | | Apraxia as sequela of ischemic cerebrovascular accident | 42535687 | Condition | SNOMED |
| | | Limb-kinetic apraxia | 4219807 | Condition | SNOMED |
| Date of Stroke | Narrower | When did the first stroke occur [Date and time] [PhenX] | 40768408 | Observation | LOINC |
| Days Post Stroke | Equivalent | Time elapsed since stroke | 36303843 | Observation | LOINC |
| First Stroke *yes/no* | N/A | Unmapped | | | |
| Motor Paresis *presence/history* | Narrower | Ataxic hemiparesis | 43530730 | Condition | SNOMED |
| | | Bilateral paresis | 4133725 | Condition | SNOMED |
| | | Cerebral hemiparesis | 4042744 | Condition | SNOMED |
| | | Cerebral paraparesis | 4231447 | Condition | SNOMED |
| | | Crossed hemiparesis | 4104652 | Condition | SNOMED |
| | | Facial hemiparesis | 4068034 | Condition | SNOMED |
| | | Functional monoparesis | 37110546 | Condition | SNOMED |
| | | Hemiparesis | 4065756 | Condition | SNOMED |
| | | Left hemiparesis | 4081504 | Condition | SNOMED |
| | | Monoparesis | 4194210 | Condition | SNOMED |
| | | Monoparesis - arm | 4093682 | Condition | SNOMED |
| | | Monoparesis of left lower limb | 37173104 | Condition | SNOMED |
| | | Monoparesis of left upper limb | 37173102 | Condition | SNOMED |

| B. Stroke-specific Medical History | | | | | |
|---|---|---|---|---|---|
| **Data Element** | **Mapping equivalency level** | **OMOP Concept Name** | **OMOP Concept ID** | **OMOP Domain** | **Source Vocabulary** |
| Motor Paresis *presence/history* *(cont.)* | Narrower *(cont.)* | Monoparesis of lower limb | 4097164 | Condition | SNOMED |
| | | Monoparesis of right lower limb | 37173100 | Condition | SNOMED |
| | | Monoparesis of right upper limb | 37173098 | Condition | SNOMED |
| | | Paraparesis | 4081615 | Condition | SNOMED |
| | | Paresis of lower extremity | 4010010 | Condition | SNOMED |
| | | Partial bilateral paresis | 4179040 | Condition | SNOMED |
| | | Right hemiparesis | 4083064 | Condition | SNOMED |
| | | Spastic paresis | 4154696 | Condition | SNOMED |
| | | Tetraparesis | 444419 | Condition | SNOMED |
| | | Triparesis | 4173242 | Condition | SNOMED |
| Stroke Type *hemorrhagic/ischemic* | N/A | Unmapped | | | |
| Stroke Chronicity | N/A | Unmapped | | | |
| TOAST Classification | Equivalent | TOAST Classification of subtypes of acute ischemic stroke | 36304812 | Observation | LOINC |
| Total Number of Strokes | N/A | Unmapped | | | |
| Unilateral Neglect *presence/history* | Equivalent | Neglect of affected side | 4078341 | Condition | SNOMED |
| | Narrower | Hemispatial neglect | 4138543 | Condition | SNOMED |
| | | Neglect of arm | 4079164 | Condition | SNOMED |
| | | Neglect of left side of body | 4113235 | Condition | SNOMED |
| | | Neglect of leg | 4086184 | Condition | SNOMED |
| | | Neglect of right side of body | 4100610 | Condition | SNOMED |
| | | Unilateral neglect [CCC] | 3002627 | Observation | LOINC |
| | | Visual neglect | 4081314 | Observation | SNOMED |

| C. Other Medical History | | | | | |
|---|---|---|---|---|---|
| **Data Element** | **Mapping equivalency level** | **OMOP Concept Name** | **OMOP Concept ID** | **OMOP Domain** | **Source Vocabulary** |
| Atrial Fibrillation (AFib) *presence/history* | Equivalent | Atrial fibrillation | 313217 | Condition | SNOMED |
| | | Atrial fibrillation | 45883018 | Meas Value | LOINC |
| | Narrower | Atrial fibrillation detected | 42689664 | Condition | SNOMED |
| | | Chronic atrial fibrillation | 4141360 | Condition | SNOMED |
| | | Controlled atrial fibrillation | 4117112 | Condition | SNOMED |
| | | ECG: atrial fibrillation | 4064452 | Condition | SNOMED |
| | | Ever told you have or had atrial fibrillation | 40761369 | Observation | LOINC |
| | | Longstanding persistent atrial fibrillation | 45768480 | Condition | SNOMED |
| | | Permanent atrial fibrillation | 4232691 | Condition | SNOMED |
| | | Persistent atrial fibrillation | 4232697 | Condition | SNOMED |
| Carotid Stenosis *presence/history* | Equivalent | Carotid artery stenosis | 442615 | Condition | SNOMED |
| | | Carotid atherosclerosis | 4102124 | Condition | SNOMED |
| | Narrower | Asymptomatic carotid artery stenosis | 761930 | Condition | SNOMED |
| | | Bilateral stenosis of carotid arteries | 37204002 | Condition | SNOMED |
| | | Internal carotid artery stenosis | 4121624 | Condition | SNOMED |
| | | Left carotid artery stenosis | 43020498 | Condition | SNOMED |
| | | Right carotid artery stenosis | 43021859 | Condition | SNOMED |
| | | Symptomatic carotid artery stenosis | 44784634 | Condition | SNOMED |

| C. Other Medical History | | | | | |
|---|---|---|---|---|---|
| **Data Element** | **Mapping equivalency level** | **OMOP Concept Name** | **OMOP Concept ID** | **OMOP Domain** | **Source Vocabulary** |
| Chronic Obstructive Pulmonary Disease (COPD) *presence/history* | Broader | Emphysema or COPD [Minimum Data Set] | 3044760 | Observation | LOINC |
| | Equivalent | COPD | 45877605 | Meas Value | LOINC |
| | | Chronic Obstructive Pulmonary Disease (COPD) | 1472264 | Meas Value | LOINC |
| | | Chronic obstructive pulmonary disease | 255573 | Condition | SNOMED |
| | | Chronic obstructive pulmonary disease | 36308250 | Meas Value | LOINC |
| Coronary Artery Disease (CAD) *presence/history* | Equivalent | Cardio/pulm: Coronary artery disease | 36309684 | Meas Value | LOINC |
| | | Coronary arteriosclerosis | 317576 | Condition | SNOMED |
| | | Coronary artery disease (CAD) (e.g., angina, myocardial infarction, and atherosclerotic heart disease (ASHD)) | 36307799 | Meas Value | LOINC |
| | | Disorder of coronary artery | 4187067 | Condition | SNOMED |
| | | History of coronary artery disease | 3168727 | Observation | Nebraska Lexicon |
| | Narrower | Coronary artery disease in last 7 days [MDSv3] | 40757908 | Observation | LOINC |
| | | Diffuse disease of coronary artery | 4162739 | Condition | SNOMED |
| | | Double coronary vessel disease | 4108673 | Condition | SNOMED |
| | | Left main coronary artery disease | 4155962 | Condition | SNOMED |
| | | Multi vessel coronary artery disease | 4155007 | Condition | SNOMED |
| | | Single coronary vessel disease | 4111393 | Condition | SNOMED |
| | | Triple vessel disease of the heart | 4124682 | Condition | SNOMED |
| Diabetes *presence/history* | Equivalent | Diabetes | 45879799 | Meas Value | LOINC |
| | | Diabetes | 35817874 | Observation | UK Biobank |
| | | Diabetes mellitus | 201820 | Condition | SNOMED |
| | | Diabetes mellitus | 45876545 | Meas Value | LOINC |
| | Narrower | Diabetes Type 1 | 45883360 | Meas Value | LOINC |
| | | Diabetes Type 2 | 45877606 | Meas Value | LOINC |
| | | Diabetes Type I | 3968674 | Meas Value | LOINC |
| | | Diabetes Type II | 3968732 | Meas Value | LOINC |
| | | Diabetes status [Identifier] | 3030608 | Observation | LOINC |
| | | History of Diabetes (regardless of treatment) [PhenX] | 40769338 | Observation | LOINC |
| High Cholesterol *presence/history* | Broader | Dyslipidemia | 4159131 | Condition | SNOMED |
| | Equivalent | Elevated cholesterol | 45883494 | Meas Value | LOINC |
| | | High Cholesterol/Hyperlipidemia | 45882058 | Meas Value | LOINC |
| | | High blood cholesterol | 45877280 | Meas Value | LOINC |
| | | Hypercholesterolemia | 4029305 | Condition | SNOMED |
| | Narrower | Ever told by doctor or nurse that you have high blood cholesterol | 40761393 | Observation | LOINC |
| Hypertension (HTN) *presence/history* | Equivalent | High blood pressure | 45883495 | Meas Value | LOINC |
| | | High blood pressure | 35819550 | Observation | UK Biobank |
| | | High blood pressure | 35819549 | Observation | UK Biobank |
| | | Hypertension | 45876622 | Meas Value | LOINC |
| | | Hypertension (high blood pressure) | 1989868 | Meas Value | LOINC |

| C. Other Medical History | | | | | |
|---|---|---|---|---|---|
| **Data Element** | **Mapping equivalency level** | **OMOP Concept Name** | **OMOP Concept ID** | **OMOP Domain** | **Source Vocabulary** |
| Hypertension (HTN) *presence/history* *(cont.)* | Equivalent *(cont.)* | Hypertension or high blood pressure | 45879470 | Meas Value | LOINC |
| | | Hypertensive disorder | 316866 | Condition | SNOMED |
| | Narrower | Diastolic hypertension | 4167358 | Condition | SNOMED |
| | | Diastolic hypertension and systolic hypertension | 42538697 | Condition | SNOMED |
| | | Essential hypertension | 320128 | Condition | SNOMED |
| | | Ever told by doctor or nurse that you have high blood pressure | 40761396 | Observation | LOINC |
| | | Hypertension stage 1 | 37311147 | Condition | SNOMED |
| | | Hypertension stage 2 | 37311148 | Condition | SNOMED |
| | | Severe hypertension (NICE - National Institute for Health and Clinical Excellence 2011) | 44809027 | Condition | SNOMED |
| | | Stage 1 hypertension (NICE - National Institute for Health and Clinical Excellence 2011) | 44809026 | Condition | SNOMED |
| | | Stage 2 hypertension (NICE - National Institute for Health and Clinical Excellence 2011) | 44809569 | Condition | SNOMED |
| | | Systolic hypertension | 4209293 | Condition | SNOMED |
| Myocardial Infarction (MI) *presence/history* | Broader | Myocardial infarction or chest pain | 45880412 | Meas Value | LOINC |
| | | Myocardial infarction, stroke | 45882239 | Meas Value | LOINC |
| | Equivalent | Cardio/pulm: Myocardial infarction | 36310882 | Meas Value | LOINC |
| | | Heart attack | 36210764 | Meas Value | LOINC |
| | | Heart attack | 35819523 | Observation | UK Biobank |
| | | Heart attack (myocardial infarction) | 36310216 | Meas Value | LOINC |
| | | Heart attack or M.I. | 45883200 | Meas Value | LOINC |
| | | Myocardial infarction | 40761359 | Observation | LOINC |
| | | Myocardial infarction | 4329847 | Condition | SNOMED |
| | | Myocardial infarction | 45881971 | Meas Value | LOINC |
| | Narrower | EKG: myocardial infarction | 4064609 | Condition | SNOMED |
| | | EKG: old myocardial infarction | 4064348 | Condition | SNOMED |
| | | Ever told by doctor that you had a myocardial infarction or heart attack | 40761439 | Observation | LOINC |
| | | History of myocardial infarction | 36309015 | Meas Value | LOINC |
| | | Myocardial infarction [Reported] | 40769284 | Observation | LOINC |
| | | Myocardial infarction in interim | 40761424 | Observation | LOINC |
| | | Myocardial infarction in past 24 hours | 21491677 | Observation | LOINC |
| | | Old myocardial infarction | 314666 | Condition | SNOMED |
| | | Previous myocardial infarction | 1620969 | Meas Value | LOINC |
| Peak Oxygen Consumption | Equivalent | Oxygen consumption (VO2) --peak | 1259612 | Measurement | LOINC |
| Peripheral Arterial Occlusive Disease *presence/history* | Broader | Ever told by doctor that you had claudication or peripheral arterial disease | 40761363 | Observation | LOINC |
| | | Peripheral arterial disease | 3654996 | Condition | SNOMED |
| | | Peripheral arterial disease | 36308312 | Meas Value | LOINC |
| | | Peripheral vascular disease (PVD) or peripheral arterial disease (PAD) | 45885065 | Meas Value | LOINC |

| C. Other Medical History | | | | | |
|---|---|---|---|---|---|
| **Data Element** | **Mapping equivalency level** | **OMOP Concept Name** | **OMOP Concept ID** | **OMOP Domain** | **Source Vocabulary** |
| Peripheral Arterial Occlusive Disease *presence/history (cont.)* | Equivalent | Peripheral arterial occlusive disease | 317309 | Condition | SNOMED |
| | Narrower | Atherosclerotic occlusive disease | 4030425 | Condition | SNOMED |
| | | Non-atherosclerotic chronic arterial occlusive disease | 37110251 | Condition | SNOMED |
| Transient Ischemic Attack (TIA) *presence/history* | Broader | Cerebrovascular accident (CVA), Transient Ischemic Attack (presence/history), or stroke | 36309963 | Meas Value | LOINC |
| | | Cerebrovascular accident/transient ischemic attack/stroke in last 7 days [MDSv3] | 40757938 | Observation | LOINC |
| | | Stroke and TIA | 44804386 | Observation | SNOMED |
| | Equivalent | Temporary stroke like symptoms (TIA - transient ischemic attack/mini-stroke) | 36308885 | Meas Value | LOINC |
| | | Transient cerebral ischemia | 373503 | Condition | SNOMED |
| | | Transient ischemic attack - TIA | 45876543 | Meas Value | LOINC |
| | Narrower | Recurrent transient cerebral ischemic attack | 40485430 | Condition | SNOMED |
| | | Transient ischemic attack [Minimum Data Set] | 3044674 | Observation | LOINC |
| | | Transient ischemic attack due to embolism | 46272244 | Condition | SNOMED |

| D. Care/Study-related Information | | | | | |
|---|---|---|---|---|---|
| **Data Element** | **Mapping equivalency level** | **OMOP Concept Name** | **OMOP Concept ID** | **OMOP Domain** | **Source Vocabulary** |
| Control Group *yes/no* | Equivalent | Control group | 44804027 | Observation | SNOMED |
| Days in Hospital | Equivalent | Duration of inpatient stay | 4084530 | Observation | SNOMED |
| Discharge Destination | Equivalent | Planned discharge destination | 44810687 | Observation | SNOMED |
| Intervention *yes/no* | Equivalent | Intervention regime | 4021163 | Procedure | SNOMED |
| | | Interventional procedure | 706394 | Meas Value | LOINC |
| | Narrower | Brief intervention | 45770015 | Procedure | SNOMED |
| | | Health intervention | 44804044 | Observation | SNOMED |
| Intervention Notes | Equivalent | Intervention synopsis | 40760317 | Observation | LOINC |
| | | Interventional procedure note | 3030648 | Note | LOINC |
| | | Interventions Narrative | 40765128 | Observation | LOINC |
| Tissue Plasminogen Activator (tPA) | Narrower | Intra-arterial injection of tissue plasminogen activator | 37158321 | Procedure | SNOMED |
| | | Intravenous injection of tissue plasminogen activator | 37158320 | Procedure | SNOMED |
| Treatment Group | N/A | Unmapped | | | |

**Supplemental Table 3.** Final mapping of assessment data elements to Observational Medical Outcomes Partnership (OMOP) standard concepts

| A. Cognitive Assessments | | | | | |
|---|---|---|---|---|---|
| **Assessment** | **Granularity Level** | **OMOP Concept Name** | **OMOP Concept ID** | **OMOP Domain** | **Source Vocabulary** |
| Aachen Aphasia Test (AAT) | Assessment-level | Aachen aphasia test | 4167592 | Measurement | SNOMED |
| | Subscale-level | Unmapped | | | |
| | Item-level | Unmapped | | | |
| Animal Naming (Category Fluency Test) | Assessment-level | Unmapped | | | |
| Benton Visual Retention Test (BVRT) | Assessment-level | Benton visual retention test | 40480548 | Measurement | SNOMED |
| | | Psychologic test, Benton visual retention test | 4148772 | Procedure | SNOMED |
| | Item-level | Unmapped | | | |
| Boston Naming Test (BNT) | Assessment-level | Boston naming test | 4164808 | Measurement | SNOMED |
| | Item-level | Unmapped | | | |
| Boston Naming Test 15-item (BNT-15) | Assessment-level | Unmapped | | | |
| | Item-level | Unmapped | | | |
| California Verbal Learning Test (CVLT) | Assessment-level | CVLT-II (California Verbal Learning Test Second Edition) | 46285857 | Measurement | SNOMED |
| | | California verbal learning test | 4151628 | Measurement | SNOMED |
| | Subscale-level | Unmapped | | | |
| | Item-level | Unmapped | | | |
| Clock Draw Test | Assessment-level | Can correctly draw all the numbers to indicate the hours of a clock [GPCOG] | 21492179 | Observation | LOINC |
| | Item-level | Unmapped | | | |
| CogState brief Battery (CBB) | Assessment-level | CogState Brief Battery | 3655544 | Measurement | SNOMED |
| | Item-level | Unmapped | | | |
| Cognitive Assessment Bedside for iPad (CABPad) | Assessment-level | Unmapped | | | |
| | Item-level | Unmapped | | | |
| Comprehensive Aphasia Test Battery | Assessment-level | Comprehensive aphasia test | 44805844 | Measurement | SNOMED |
| | Subscale-level | Unmapped | | | |
| | Item-level | Unmapped | | | |
| Controlled Word Association Test (COWAT; F, A, S) | Assessment-level | Unmapped | | | |
| | Item-level | Unmapped | | | |
| Corsi Block-Tapping Test | Assessment-level | Unmapped | | | |
| | Item-level | Unmapped | | | |
| Hopkins Verbal Learning Test (HVLT) | Assessment-level | Unmapped | | | |
| | Subscale-level | Unmapped | | | |
| | Item-level | Unmapped | | | |
| Line Cancellation Test | Assessment-level | Unmapped | | | |
| Mehrfachwahl-Wortschatz Test (MWT) | Assessment-level | Unmapped | | | |
| | Item-level | Unmapped | | | |
| Mini Mental State Examination (MMSE) | Assessment-level | Mini-Mental State Examination [MMSE] | 42869861 | Observation | LOINC |
| | | Mini-mental state examination | 4169175 | Measurement | SNOMED |
| | | Total score [MMSE] | 42869860 | Observation | LOINC |
| | Subscale-level | Unmapped | | | |
| | Item-level | Unmapped | | | |

| A. Cognitive Assessments | | | | | |
|---|---|---|---|---|---|
| **Assessment** | **Granularity Level** | **OMOP Concept Name** | **OMOP Concept ID** | **OMOP Domain** | **Source Vocabulary** |
| Montreal Cognitive Assessment (MoCA) | Assessment-level | Montreal Cognitive Assessment [MoCA] | 43054876 | Observation | LOINC |
| | | Montreal Cognitive Assessment scale | 3187793 | Measurement | Nebraska Lexicon |
| | | Montreal Cognitive Assessment version 7.1 | 606673 | Measurement | SNOMED |
| | | Montreal Cognitive Assessment version 7.2 | 606672 | Measurement | SNOMED |
| | | Montreal Cognitive Assessment version 7.3 | 606670 | Measurement | SNOMED |
| | | Montreal Cognitive Assessment version 8.1 | 606671 | Measurement | SNOMED |
| | | Montreal cognitive assessment | 44808666 | Measurement | SNOMED |
| | | Total score [MoCA] | 43054915 | Observation | LOINC |
| | Subscale-level | Unmapped | | | |
| | Item-level | Unmapped | | | |
| Neuro-QoL Cognitive Function | Assessment-level | Unmapped | | | |
| | Item-level | How much difficulty do you currently have learning new tasks or instructions [NeuroQol] | 40770556 | Observation | LOINC |
| | | How much difficulty do you currently have managing your time to do most of your daily activities [NeuroQol] | 40770553 | Observation | LOINC |
| | | How much difficulty do you currently have planning for and keeping appointments that are not part of your weekly routine, like a therapy or doctor appointment, or a social gathering with friends and family [NeuroQol] | 40770552 | Observation | LOINC |
| | | How much difficulty do you currently have reading and following complex instructions, like directions for a new medication [NeuroQol] | 40770551 | Observation | LOINC |
| | | I had to read something several times to understand it in the past 7 days [NeuroQol] | 40770562 | Observation | LOINC |
| | | I had to work really hard to pay attention or I would make a mistake in the past 7 days [NeuroQol] | 40770568 | Observation | LOINC |
| | | I had trouble concentrating in the past 7 days [NeuroQol] | 40770569 | Observation | LOINC |
| | | My thinking was slow in the past 7 days [NeuroQol] | 40770567 | Observation | LOINC |
| NeuroCogFX | Assessment-level | Unmapped | | | |
| | Subscale-level | Unmapped | | | |
| | Item-level | Unmapped | | | |
| Phonological/Phonemic Fluency Test (P, M, R) | Assessment-level | Unmapped | | | |
| | Item-level | Unmapped | | | |
| Repeatable Battery for the Assessment of Neuropsychological Status (RBANS) | Assessment-level | Repeatable battery for the assessment of neuropsychological status | 44802487 | Measurement | SNOMED |
| | Subscale-level | Unmapped | | | |
| | Item-level | Unmapped | | | |

**A. Cognitive Assessments**

| Assessment | Granularity Level | OMOP Concept Name | OMOP Concept ID | OMOP Domain | Source Vocabulary |
|---|---|---|---|---|---|
| Rey Auditory Verbal Learning Test (RAVLT) | Assessment-level | Unmapped | | | |
| | Subscale-level | Unmapped | | | |
| | Item-level | Unmapped | | | |
| Rey-Osterrieth Complex Figure Test | Assessment-level | Rey complex figure test | 4208363 | Measurement | SNOMED |
| | | Rey figure test | 4145250 | Measurement | SNOMED |
| | Subscale-level | Unmapped | | | |
| | Item-level | Unmapped | | | |
| Semantic Verbal Fluency Test | Assessment-level | Unmapped | | | |
| Short Blessed Test (SBT) | Assessment-level | Unmapped | | | |
| | Item-level | Unmapped | | | |
| Star Cancellation Task | Assessment-level | Unmapped | | | |
| Stroop Color and Word Test | Assessment-level | Stroop neuropsychological screening test | 4136943 | Measurement | SNOMED |
| | Item-level | Stroop test: number of errors in first subtask | 738835 | Measurement | OMOP Extension |
| | | Stroop test: number of errors in second subtask | 738845 | Measurement | OMOP Extension |
| | | Stroop test: number of errors in third subtask | 738826 | Measurement | OMOP Extension |
| | | Stroop test: time of first subtask | 738832 | Measurement | OMOP Extension |
| | | Stroop test: time of second subtask | 738870 | Measurement | OMOP Extension |
| | | Stroop test: time of third subtask | 738833 | Measurement | OMOP Extension |
| Symbol Digit Modalities Test / Digit Symbol Substitution Task | Assessment-level | Symbol digit modalities test | 4166621 | Measurement | SNOMED |
| | Item-level | Unmapped | | | |
| Test of Premorbid Function (TOPF) | Assessment-level | Unmapped | | | |
| Token Test (16-item) | Assessment-level | Revised token test | 4121523 | Measurement | SNOMED |
| | | Token test | 4169492 | Measurement | SNOMED |
| | Item-level | Unmapped | | | |
| Trail Making Test (TMT) | Assessment-level | Trail making test | 4165596 | Measurement | SNOMED |
| | Item-level | Unmapped | | | |
| Wechsler Adult Intelligence Scale (WAIS) | Assessment-level | Intelligence test/WAIS | 4241571 | Procedure | SNOMED |
| | | Wechsler Adult Intelligence Scale 4th Edition (WAIS-IV) | 40219612 | Measurement | OMOP Extension |
| | | Wechsler Adult Intelligence Scale fourth edition | 37168546 | Measurement | SNOMED |
| | | Wechsler adult intelligence scale | 4165604 | Measurement | SNOMED |
| | | Wechsler adult intelligence scale - revised | 4166746 | Measurement | SNOMED |
| | Subscale-level | WAIS-IV: Full Scale IQ (FSIQ) | 40219539 | Measurement | OMOP Extension |
| | | WAIS-IV: General Ability Index (GAI) | 40219509 | Measurement | OMOP Extension |
| | | WAIS-IV: Perceptual Reasoning Index (PRI) | 40219497 | Measurement | OMOP Extension |
| | | WAIS-IV: Processing Speed Index (PSI) | 40219545 | Measurement | OMOP Extension |

| A. Cognitive Assessments | | | | | |
|---|---|---|---|---|---|
| **Assessment** | **Granularity Level** | **OMOP Concept Name** | **OMOP Concept ID** | **OMOP Domain** | **Source Vocabulary** |
| Wechsler Adult Intelligence Scale (WAIS) *(cont.)* | Subscale-level *(cont.)* | WAIS-IV: Verbal Comprehension Index (VCI) | 40219608 | Measurement | OMOP Extension |
| | | WAIS-IV: Working Memory Index (WMI) | 40219432 | Measurement | OMOP Extension |
| | Item-level | WAIS-IV: Perceptual Reasoning Index (PRI) - Block Design | 40219567 | Measurement | OMOP Extension |
| | | WAIS-IV: Perceptual Reasoning Index (PRI) - Figure Weights | 40219527 | Measurement | OMOP Extension |
| | | WAIS-IV: Perceptual Reasoning Index (PRI) - Matrix Reasoning | 40219500 | Measurement | OMOP Extension |
| | | WAIS-IV: Perceptual Reasoning Index (PRI) - Picture Completion | 40219457 | Measurement | OMOP Extension |
| | | WAIS-IV: Perceptual Reasoning Index (PRI) - Visual Puzzles | 40219593 | Measurement | OMOP Extension |
| | | WAIS-IV: Processing Speed Index (PSI) - Cancellation | 40219456 | Measurement | OMOP Extension |
| | | WAIS-IV: Processing Speed Index (PSI) - Coding | 40219515 | Measurement | OMOP Extension |
| | | WAIS-IV: Processing Speed Index (PSI) - Symbol Search | 40219538 | Measurement | OMOP Extension |
| | | WAIS-IV: Verbal Comprehension Index (VCI) - Comprehension | 40219501 | Measurement | OMOP Extension |
| | | WAIS-IV: Verbal Comprehension Index (VCI) - Information | 40219601 | Measurement | OMOP Extension |
| | | WAIS-IV: Verbal Comprehension Index (VCI) - Similarities | 40219426 | Measurement | OMOP Extension |
| | | WAIS-IV: Verbal Comprehension Index (VCI) - Vocabulary | 40219512 | Measurement | OMOP Extension |
| | | WAIS-IV: Working Memory Index (WMI) - Arithmetic | 40219525 | Measurement | OMOP Extension |
| | | WAIS-IV: Working Memory Index (WMI) - Digit span | 40219496 | Measurement | OMOP Extension |
| | | WAIS-IV: Working Memory Index (WMI) - Letter-Number Sequencing | 40219558 | Measurement | OMOP Extension |

| B. Global Function Assessments | | | | | |
|---|---|---|---|---|---|
| **Assessment** | **Granularity Level** | **OMOP Concept Name** | **OMOP Concept ID** | **OMOP Domain** | **Source Vocabulary** |
| Barthel Index / Modified Barthel Index (BI/MBI) | Assessment-level | Barthel Index of Activities of Daily Living score | 37117906 | Measurement | SNOMED |
| | | Barthel index | 4167605 | Measurement | SNOMED |
| | | Barthel index panel | 1617262 | Measurement | LOINC |
| | | Barthel original index of activities of daily living score | 44802296 | Measurement | SNOMED |
| | | Modified Barthel index of activities of daily living | 40481416 | Measurement | SNOMED |
| | | Total score Barthel Index | 1617389 | Measurement | LOINC |
| | Item-level | Ambulation - functional ability | 42529380 | Observation | LOINC |
| | | Bathing - functional ability | 42529375 | Observation | LOINC |
| | | Bladder control - functional ability | 1617085 | Observation | LOINC |
| | | Bowel control - functional ability | 1617512 | Observation | LOINC |
| | | Dressing - functional ability | 42529376 | Observation | LOINC |
| | | Feeding or eating - functional ability | 42529378 | Observation | LOINC |
| | | Grooming - functional ability | 1616957 | Observation | LOINC |

| B. Global Function Assessments | | | | | |
|---|---|---|---|---|---|
| **Assessment** | **Granularity Level** | **OMOP Concept Name** | **OMOP Concept ID** | **OMOP Domain** | **Source Vocabulary** |
| Barthel Index / Modified Barthel Index (BI/MBI) *(cont.)* | Item-level *(cont.)* | Stairs - functional ability | 1616510 | Observation | LOINC |
| | | Toileting - functional ability | 42529377 | Observation | LOINC |
| | | Transferring - functional ability | 42529379 | Observation | LOINC |
| Bogenhausen Dysphagia Score (BODS) | Assessment-level | Unmapped | | | |
| | Subscale-level | Unmapped | | | |
| | Item-level | Unmapped | | | |
| Chalder Fatigue Scale (CFQ-11) | Assessment-level | Chalder Fatigue Scale | 42690519 | Measurement | SNOMED |
| Chalder Fatigue Scale (CFQ-11) | Subscale-level | Unmapped | | | |
| Chalder Fatigue Scale (CFQ-11) | Item-level | Unmapped | | | |
| Charlson Comorbidity Index (CCI) | Assessment-level | Age-adjusted Charlson comorbidity Index | 715970 | Measurement | OMOP Extension |
| Charlson Comorbidity Index (CCI) | Assessment-level | Charlson Comorbidity Index | 42538860 | Measurement | SNOMED |
| Charlson Comorbidity Index (CCI) | Item-level | Unmapped | | | |
| EuroQoL Five Dimension Five Level Questionnaire (EQ-5D-5L) | Assessment-level | EuroQoL five dimension five level questionnaire | 37159771 | Measurement | SNOMED |
| | Assessment-level | EuroQoL five dimension questionnaire | 40483273 | Measurement | SNOMED |
| | Assessment-level | EuroQol five dimension five level index value | 42537273 | Observation | SNOMED |
| | Assessment-level | EuroQol five dimension five level scale | 44806410 | Measurement | SNOMED |
| | Assessment-level | EuroQol five dimension self-report questionnaire | 44807984 | Measurement | SNOMED |
| | Item-level | EuroQol five dimension five level anxiety depression score | 44813556 | Measurement | SNOMED |
| | Item-level | EuroQol five dimension five level mobility score | 44806412 | Measurement | SNOMED |
| | Item-level | EuroQol five dimension five level pain discomfort score | 44806414 | Measurement | SNOMED |
| | Item-level | EuroQol five dimension five level self-care score | 44806413 | Measurement | SNOMED |
| | Item-level | EuroQol five dimension five level usual activities score | 44813555 | Measurement | SNOMED |
| Fatigue Severity Scale (FSS) | Assessment-level | Unmapped | | | |
| | Subscale-level | Unmapped | | | |
| | Item-level | Unmapped | | | |
| Fugl-Meyer Assessment (FMA) All-domain Score | Assessment-level | Unmapped | | | |
| IRF-PAI Mobility | Assessment-level | IRF-PAI - Mobility - admission performance during 3 day assessment period [CMS Assessment] | 36204569 | Observation | LOINC |
| | | IRF-PAI - Mobility - discharge performance during 3 day assessment period [CMS Assessment] | 36204577 | Observation | LOINC |
| | Item-level | 1 step (curb) - functional ability during 3 day assessment period [CMS Assessment] | 36204502 | Observation | LOINC |

| B. Global Function Assessments | | | | | |
|---|---|---|---|---|---|
| **Assessment** | **Granularity Level** | **OMOP Concept Name** | **OMOP Concept ID** | **OMOP Domain** | **Source Vocabulary** |
| IRF-PAI Mobility *(cont.)* | Item-level *(cont.)* | 1 step (curb) - functional ability during assessment period [CMS Assessment] | 36305669 | Observation | LOINC |
| | | 12 steps - functional ability during 3 day assessment period [CMS Assessment] | 36204498 | Observation | LOINC |
| | | 12 steps - functional ability during assessment period [CMS Assessment] | 36306094 | Observation | LOINC |
| | | 4 steps - functional ability during 3 day assessment period [CMS Assessment] | 36204500 | Observation | LOINC |
| | | 4 steps - functional ability during assessment period [CMS Assessment] | 36304093 | Observation | LOINC |
| | | Bed-to-chair transfer - functional ability during 3 day assessment period [CMS Assessment] | 36204516 | Observation | LOINC |
| | | Bed-to-chair transfer - functional ability during assessment period [CMS Assessment] | 36305770 | Observation | LOINC |
| | | Car transfer - functional ability during 3 day assessment period [CMS Assessment] | 36204512 | Observation | LOINC |
| | | Car transfer - functional ability during assessment period [CMS Assessment] | 36305143 | Observation | LOINC |
| | | Does the patient use a wheelchair or scooter during 3 day assessment period [CMS Assessment] | 36204571 | Observation | LOINC |
| | | Does the patient use a wheelchair/scooter during assessment period [CMS Assessment] | 36031452 | Observation | LOINC |
| | | Does the patient walk - admission performance during 3 day assessment period [CMS Assessment] | 36204570 | Observation | LOINC |
| | | Does the patient walk - discharge performance during 3 day assessment period [CMS Assessment] | 36204578 | Observation | LOINC |
| | | Does the patient walk during assessment period [CMS Assessment] | 36659958 | Observation | LOINC |
| | | Indicate the type of wheelchair or scooter used during 3 day assessment period [CMS Assessment] | 36204572 | Observation | LOINC |
| | | Indicate the type of wheelchair/scooter used during assessment period [CMS Assessment] | 36031511 | Observation | LOINC |
| | | Lying to sitting on side of bed - functional ability during 3 day assessment period [CMS Assessment] | 36204520 | Observation | LOINC |

| B. Global Function Assessments | | | | | |
|---|---|---|---|---|---|
| **Assessment** | **Granularity Level** | **OMOP Concept Name** | **OMOP Concept ID** | **OMOP Domain** | **Source Vocabulary** |
| IRF-PAI Mobility *(cont.)* | Item-level *(cont.)* | Lying to sitting on side of bed - functional ability during assessment period [CMS Assessment] | 36305499 | Observation | LOINC |
| | | Lying to sitting on side of bed - most dependent performance during the past month [CMS Assessment] | 36660324 | Observation | LOINC |
| | | Lying to sitting on side of bed - usual functional ability during assessment period [CMS Assessment] | 36660567 | Observation | LOINC |
| | | Picking up object - functional ability during 3 day assessment period [CMS Assessment] | 36204496 | Observation | LOINC |
| | | Picking up object - functional ability during assessment period [CMS Assessment] | 36305358 | Observation | LOINC |
| | | Picking up object - usual functional ability during assessment period [CMS Assessment] | 36660137 | Observation | LOINC |
| | | Roll left and right - functional ability during 3 day assessment period [CMS Assessment] | 36204524 | Observation | LOINC |
| | | Roll left and right - functional ability during assessment period [CMS Assessment] | 36303535 | Observation | LOINC |
| | | Roll left and right - usual functional ability during assessment period [CMS Assessment] | 36660309 | Observation | LOINC |
| | | Sit to lying - functional ability during 3 day assessment period [CMS Assessment] | 36204522 | Observation | LOINC |
| | | Sit to lying - functional ability during assessment period [CMS Assessment] | 36303964 | Observation | LOINC |
| | | Sit to lying - usual functional ability during assessment period [CMS Assessment] | 36659724 | Observation | LOINC |
| | | Sit to stand - functional ability during 3 day assessment period [CMS Assessment] | 36204518 | Observation | LOINC |
| | | Sit to stand - functional ability during assessment period [CMS Assessment] | 36306060 | Observation | LOINC |
| | | Sit to stand - usual functional ability during assessment period [CMS Assessment] | 36660351 | Observation | LOINC |
| | | Toilet transfer - functional ability during 3 day assessment period [CMS Assessment] | 36204514 | Observation | LOINC |
| | | Toilet transfer - usual functional ability during assessment period [CMS Assessment] | 36660359 | Observation | LOINC |
| | | Toilet transferring - functional ability during assessment period [CMS Assessment] | 40760363 | Observation | LOINC |
| | | Walk 10 feet - functional ability during 3 day assessment period [CMS Assessment] | 36204510 | Observation | LOINC |

| B. Global Function Assessments | | | | | |
|---|---|---|---|---|---|
| **Assessment** | **Granularity Level** | **OMOP Concept Name** | **OMOP Concept ID** | **OMOP Domain** | **Source Vocabulary** |
| IRF-PAI Mobility *(cont.)* | Item-level *(cont.)* | Walk 10 feet - functional ability during assessment period [CMS Assessment] | 36305711 | Observation | LOINC |
| | | Walk 10 feet - usual functional ability during assessment period [CMS Assessment] | 36660254 | Observation | LOINC |
| | | Walk 150 feet - functional ability during 3 day assessment period [CMS Assessment] | 36204506 | Observation | LOINC |
| | | Walk 150 feet - functional ability during assessment period [CMS Assessment] | 36304861 | Observation | LOINC |
| | | Walk 150 feet - usual functional ability during assessment period [CMS Assessment] | 36659785 | Observation | LOINC |
| | | Walk 50 feet with two turns - functional ability during 3 day assessment period [CMS Assessment] | 36204508 | Observation | LOINC |
| | | Walk 50 feet with two turns - functional ability during assessment period [CMS Assessment] | 36304179 | Observation | LOINC |
| | | Walk 50 feet with two turns - usual functional ability during assessment period [CMS Assessment] | 36659711 | Observation | LOINC |
| | | Walking 10 feet on uneven surfaces - functional ability during 3 day assessment period [CMS Assessment] | 36204504 | Observation | LOINC |
| | | Walking 10 feet on uneven surfaces - functional ability during assessment period [CMS Assessment] | 36304903 | Observation | LOINC |
| | | Walking 10 feet on uneven surfaces - usual functional ability during assessment period [CMS Assessment] | 36659731 | Observation | LOINC |
| | | Wheel 150 feet - functional ability during 3 day assessment period [CMS Assessment] | 36204541 | Observation | LOINC |
| | | Wheel 150 feet - functional ability during assessment period [CMS Assessment] | 36305478 | Observation | LOINC |
| | | Wheel 150 feet - usual functional ability during assessment period [CMS Assessment] | 36659802 | Observation | LOINC |
| | | Wheel 50 feet with two turns - functional ability during 3 day assessment period [CMS Assessment] | 36204494 | Observation | LOINC |
| | | Wheel 50 feet with two turns - functional ability during assessment period [CMS Assessment] | 36303335 | Observation | LOINC |
| | | Wheel 50 feet with two turns - usual functional ability during assessment period [CMS Assessment] | 36660010 | Observation | LOINC |

| B. Global Function Assessments | | | | | |
|---|---|---|---|---|---|
| **Assessment** | **Granularity Level** | **OMOP Concept Name** | **OMOP Concept ID** | **OMOP Domain** | **Source Vocabulary** |
| IRF-PAI Self-Care | Assessment-level | IRF-PAI v3.0, MDS v1.17.1, 1.17.2 - Self-care - admission performance during assessment period [CMS Assessment] | 36031225 | Observation | LOINC |
| | | IRF-PAIv3.0, MDS v1.17.1, 1.17.2 - Self-care - discharge performance during assessment period [CMS Assessment] | 36031304 | Observation | LOINC |
| | | Self-care - admission performance [CMS Assessment] | 36204539 | Observation | LOINC |
| | | Self-care - discharge performance [CMS Assessment] | 36204559 | Observation | LOINC |
| | Item-level | Eating - functional ability during 3 day assessment period [CMS Assessment] | 36204538 | Observation | LOINC |
| | | Eating - functional ability during assessment period [CMS Assessment] | 36305354 | Observation | LOINC |
| | | Eating - usual functional ability during assessment period [CMS Assessment] | 36660604 | Observation | LOINC |
| | | Lower body dressing - functional ability during 3 day assessment period [CMS Assessment] | 36204528 | Observation | LOINC |
| | | Lower body dressing - functional ability during assessment period [CMS Assessment] | 36305884 | Observation | LOINC |
| | | Lower body dressing - usual functional ability during assessment period [CMS Assessment] | 36660409 | Observation | LOINC |
| | | Oral hygiene - functional ability during 3 day assessment period [CMS Assessment] | 36204536 | Observation | LOINC |
| | | Oral hygiene - functional ability during assessment period [CMS Assessment] | 36306183 | Observation | LOINC |
| | | Oral hygiene - usual functional ability during assessment period [CMS Assessment] | 36660465 | Observation | LOINC |
| | | Personal hygiene - self-performance during assessment period [CMS Assessment] | 3043616 | Observation | LOINC |
| | | Putting on and taking off footwear - functional ability during assessment period [CMS Assessment] | 36303908 | Observation | LOINC |
| | | Putting on/taking off footwear - functional ability during 3 day assessment period [CMS Assessment] | 36204526 | Observation | LOINC |
| | | Putting on/taking off footwear - usual functional ability during assessment period [CMS Assessment] | 36660380 | Observation | LOINC |
| | | Shower/bathe self - functional ability during 3 day assessment period [CMS Assessment] | 36204532 | Observation | LOINC |

**B. Global Function Assessments**

| Assessment | Granularity Level | OMOP Concept Name | OMOP Concept ID | OMOP Domain | Source Vocabulary |
|---|---|---|---|---|---|
| IRF-PAI Self-Care *(cont.)* | Item-level *(cont.)* | Shower/bathe self - functional ability during assessment period [CMS Assessment] | 36303469 | Observation | LOINC |
| | | Shower/bathe self - usual functional ability during assessment period [CMS Assessment] | 36659730 | Observation | LOINC |
| | | Toileting hygiene - functional ability during 3 day assessment period [CMS Assessment] | 36204534 | Observation | LOINC |
| | | Toileting hygiene - functional ability during assessment period [CMS Assessment] | 40760364 | Observation | LOINC |
| | | Toileting hygiene - usual functional ability during assessment period [CMS Assessment] | 36659844 | Observation | LOINC |
| | | Upper body dressing - functional ability during 3 day assessment period [CMS Assessment] | 36204530 | Observation | LOINC |
| | | Upper body dressing - functional ability during assessment period [CMS Assessment] | 36305561 | Observation | LOINC |
| | | Upper body dressing - usual functional ability during assessment period [CMS Assessment] | 36660419 | Observation | LOINC |
| NIH Stroke Scale (NIHSS) | Assessment-level | NIH Stroke Scale | 42868656 | Observation | LOINC |
| | | NIH stroke scale (NIHSS) | 45884794 | Meas Value | LOINC |
| | | National Institutes of Health stroke scale | 42872749 | Measurement | SNOMED |
| | | Total score [NIH Stroke Scale] | 42869844 | Observation | LOINC |
| | Item-level | Best gaze [NIH Stroke Scale] | 42868661 | Observation | LOINC |
| | | Best language [NIH Stroke Scale] | 42868668 | Observation | LOINC |
| | | Dysarthria [NIH Stroke Scale] | 42868669 | Observation | LOINC |
| | | Extinction and inattention [NIH Stroke Scale] | 42868670 | Observation | LOINC |
| | | Facial palsy [NIH Stroke Scale] | 42868663 | Observation | LOINC |
| | | LOC commands [NIH Stroke Scale] | 42868660 | Observation | LOINC |
| | | LOC questions [NIH Stroke Scale] | 42868659 | Observation | LOINC |
| | | Level of consciousness [NIH Stroke Scale] | 42868658 | Observation | LOINC |
| | | Limb ataxia [NIH Stroke Scale] | 42868666 | Observation | LOINC |
| | | Motor arm Left arm [NIH Stroke Scale] | 42868664 | Observation | LOINC |
| | | Motor arm Right arm [NIH Stroke Scale] | 42869911 | Observation | LOINC |
| | | Motor leg Leg - left [NIH Stroke Scale] | 42868665 | Observation | LOINC |
| | | Motor leg Leg - right [NIH Stroke Scale] | 42869912 | Observation | LOINC |
| | | Sensory [NIH Stroke Scale] | 42868667 | Observation | LOINC |
| | | Visual [NIH Stroke Scale] | 42868662 | Observation | LOINC |
| Pittsburgh Sleep Quality Index (PSQI) | Assessment-level | Pittsburgh sleep quality index | 44783153 | Measurement | SNOMED |
| | Subscale-level | Unmapped | | | |
| | Item-level | Unmapped | | | |

| B. Global Function Assessments | | | | | |
|---|---|---|---|---|---|
| **Assessment** | **Granularity Level** | **OMOP Concept Name** | **OMOP Concept ID** | **OMOP Domain** | **Source Vocabulary** |
| Rankin Scale (RS) / Modified Rankin Scale (mRS) | Assessment-level | Modified Rankin Scale | 3654838 | Measurement | SNOMED |
| | | Modified rankin scale | 46235672 | Measurement | LOINC |
| | | Rankin scale | 4165307 | Measurement | SNOMED |
| Stroke Impact Scale (SIS) | Assessment-level | Stroke impact scale version 3.0 | 45767552 | Measurement | SNOMED |
| | Subscale-level | Stroke impact scale version 3.0 activities of daily living score | 45768969 | Measurement | SNOMED |
| | | Stroke impact scale version 3.0 communication score | 45768972 | Measurement | SNOMED |
| | | Stroke impact scale version 3.0 emotion score | 45768970 | Measurement | SNOMED |
| | | Stroke impact scale version 3.0 hand function score | 45768967 | Measurement | SNOMED |
| | | Stroke impact scale version 3.0 memory score | 45768971 | Measurement | SNOMED |
| | | Stroke impact scale version 3.0 mobility score | 45768968 | Measurement | SNOMED |
| | | Stroke impact scale version 3.0 physical domain score | 45771024 | Measurement | SNOMED |
| | | Stroke impact scale version 3.0 social participation score | 45768973 | Measurement | SNOMED |
| | | Stroke impact scale version 3.0 strength score | 45768966 | Measurement | SNOMED |
| | Item-level | Unmapped | | | |
| Stroke Impact Scale 16-item (SIS-16) | Assessment-level | Unmapped | | | |
| | Item-level | Unmapped | | | |
| Stroke Self Efficacy Questionnaire (SSEQ) | Assessment-level | Unmapped | | | |
| | Subscale-level | Unmapped | | | |
| | Item-level | Unmapped | | | |
| Stroke Specific Quality of Life Scale (SSQoL) | Assessment-level | Unmapped | | | |
| | Subscale-level | Unmapped | | | |
| | Item-level | Unmapped | | | |

| C. Psychosocial Assessments | | | | | |
|---|---|---|---|---|---|
| **Assessment** | **Granularity Level** | **OMOP Concept Name** | **OMOP Concept ID** | **OMOP Domain** | **Source Vocabulary** |
| Beck Depression Inventory (BDI) | Assessment-level | Beck Depression Inventory II | 1447419 | Measurement | SNOMED |
| | | Beck Depression Inventory II [BDI] | 36304859 | Observation | LOINC |
| | | Beck Depression Inventory II total score [BDI] | 36305191 | Observation | LOINC |
| | | Beck depression inventory | 4167608 | Measurement | SNOMED |
| | Item-level | Unmapped | | | |
| Center for Epidemiological Studies Depression Scale (CES-D) | Assessment-level | Center for Epidemiologic Studies Depression Scale panel [CES-D] | 1761569 | Observation | LOINC |
| | | Center for Epidemiologic Studies Depression Scale-Revised [CESD-R] | 36305791 | Observation | LOINC |
| | | Center for Epidemiologic Studies Depression Scale-Revised total score [CESD-R] | 36303297 | Observation | LOINC |
| | Subscale-level | Unmapped | | | |
| | Item-level | Bothered by things that are not usually bothersome | 1761605 | Observation | LOINC |
| | | Crying spells | 1761710 | Observation | LOINC |
| | | Enjoying life | 1761535 | Observation | LOINC |

| C. Psychosocial Assessments | | | | | |
|---|---|---|---|---|---|
| **Assessment** | **Granularity Level** | **OMOP Concept Name** | **OMOP Concept ID** | **OMOP Domain** | **Source Vocabulary** |
| Center for Epidemiological Studies Depression Scale (CES-D) *(cont.)* | Item-level *(cont.)* | Feeling depressed | 1761895 | Observation | LOINC |
| | | Feeling everything is too much of an effort | 1761503 | Observation | LOINC |
| | | Feeling fearful | 1761739 | Observation | LOINC |
| | | Feeling happy | 1761700 | Observation | LOINC |
| | | Feeling hopeful about the future | 1761675 | Observation | LOINC |
| | | Feeling just as good as others | 1761886 | Observation | LOINC |
| | | Feeling life had been a failure | 1761350 | Observation | LOINC |
| | | Feeling lonely | 1761319 | Observation | LOINC |
| | | Feeling people dislike me | 1761541 | Observation | LOINC |
| | | Feeling sad | 1761730 | Observation | LOINC |
| | | I was bothered by things that usually don't bother me [CES-DC] | 40768157 | Observation | LOINC |
| | | Lack of energy | 1761073 | Observation | LOINC |
| | | People were unfriendly | 1761353 | Observation | LOINC |
| | | Poor appetite | 1761326 | Observation | LOINC |
| | | Quieter than usual | 1761653 | Observation | LOINC |
| | | Restless sleep | 1761695 | Observation | LOINC |
| | | Trouble focusing | 1761791 | Observation | LOINC |
| | | Unhappy even with help from my family or friends | 1761611 | Observation | LOINC |
| Depression Anxiety Stress Scale 21-item (DAS-21) | Assessment-level | Unmapped | | | |
| | Subscale-level | Depression anxiety stress scales anxiety score | 4216757 | Measurement | SNOMED |
| | | Depression anxiety stress scales depression score | 4220306 | Measurement | SNOMED |
| | | Depression anxiety stress scales stress score | 4220144 | Measurement | SNOMED |
| | Item-level | Unmapped | | | |
| Frontal Behavioral Inventory (FBI) | Assessment-level | Unmapped | | | |
| | Subscale-level | Unmapped | | | |
| | Item-level | Unmapped | | | |
| Generalized Anxiety Disorder 7-item (GAD-7) | Assessment-level | Generalized Anxiety Disorder 7 item scale | 45772733 | Measurement | SNOMED |
| | | Generalized anxiety disorder 7 item (GAD-7) | 40772159 | Observation | LOINC |
| | | Generalized anxiety disorder 7 item (GAD-7) total score [Reported.PHQ] | 42868746 | Observation | LOINC |
| | Item-level | Becoming easily annoyed or irritable in last 4 weeks [Reported.PHQ] | 40772115 | Observation | LOINC |
| | | Being so restless that it is hard to sit still in last 2 weeks [Reported.PHQ] | 40772157 | Observation | LOINC |
| | | Feeling afraid as if something awful might happen in last 2 weeks [Reported.PHQ] | 40772158 | Observation | LOINC |
| | | Feeling nervous, anxious or on edge in last 2 weeks | 40772149 | Observation | LOINC |
| | | Not able to stop or control worrying in the last 2 weeks | 40771094 | Observation | LOINC |
| | | Trouble relaxing in last 2 weeks [Reported.PHQ] | 40772156 | Observation | LOINC |

| C. Psychosocial Assessments | | | | | |
|---|---|---|---|---|---|
| **Assessment** | **Granularity Level** | **OMOP Concept Name** | **OMOP Concept ID** | **OMOP Domain** | **Source Vocabulary** |
| Generalized Anxiety Disorder 7-item (GAD-7) *(cont.)* | Item-level *(cont.)* | Worrying too much about different things in last 2 weeks [Reported.PHQ] | 40772155 | Observation | LOINC |
| Geriatric Depression Scale (GDS) | Assessment-level | Geriatric depression scale | 4159706 | Measurement | SNOMED |
| | | Geriatric depression scale (GDS) panel | 3049169 | Observation | LOINC |
| | | Geriatric depression scale (GDS) total | 3051694 | Observation | LOINC |
| | | Geriatric depression scale original long form | 40482007 | Measurement | SNOMED |
| | Item-level | Are you afraid that something bad is going to happen to you [GDS] | 3052621 | Observation | LOINC |
| | | Are you basically satisfied with your life [GDS] | 3048479 | Observation | LOINC |
| | | Are you bothered by thoughts you cannot get out of your head [GDS] | 3052037 | Observation | LOINC |
| | | Are you hopeful about the future [GDS] | 3051068 | Observation | LOINC |
| | | Are you in good spirits most of the time [GDS] | 3049130 | Observation | LOINC |
| | | Do you enjoy getting up in the morning [GDS] | 3053217 | Observation | LOINC |
| | | Do you feel full of energy [GDS] | 3050101 | Observation | LOINC |
| | | Do you feel happy most of the time [GDS] | 3048472 | Observation | LOINC |
| | | Do you feel pretty worthless the way you are now [GDS] | 3051419 | Observation | LOINC |
| | | Do you feel that your life is empty [GDS] | 3052362 | Observation | LOINC |
| | | Do you feel that your situation is hopeless [GDS] | 3048841 | Observation | LOINC |
| | | Do you feel you have more problems with memory than most [GDS] | 3052630 | Observation | LOINC |
| | | Do you find life very exciting [GDS] | 3052672 | Observation | LOINC |
| | | Do you frequently feel like crying [GDS] | 3050778 | Observation | LOINC |
| | | Do you frequently get upset over little things [GDS] | 3052333 | Observation | LOINC |
| | | Do you frequently worry about the future [GDS] | 3053240 | Observation | LOINC |
| | | Do you have trouble concentrating [GDS] | 3049484 | Observation | LOINC |
| | | Do you often feel helpless [GDS] | 3051362 | Observation | LOINC |
| | | Do you often get bored [GDS] | 3048797 | Observation | LOINC |
| | | Do you often get restless and fidgety [GDS] | 3052376 | Observation | LOINC |
| | | Do you prefer to avoid social gatherings [GDS] | 3051736 | Observation | LOINC |
| | | Do you prefer to stay at home, rather than going out and doing new things [GDS] | 3049156 | Observation | LOINC |
| | | Do you think it is wonderful to be alive now [GDS] | 3051716 | Observation | LOINC |
| | | Do you think that most people are better off than you are [GDS] | 3053256 | Observation | LOINC |

| C. Psychosocial Assessments | | | | | |
|---|---|---|---|---|---|
| **Assessment** | **Granularity Level** | **OMOP Concept Name** | **OMOP Concept ID** | **OMOP Domain** | **Source Vocabulary** |
| Geriatric Depression Scale (GDS) *(cont.)* | Item-level *(cont.)* | Do you worry a lot about the past [GDS] | 3051056 | Observation | LOINC |
| | | Have you dropped many of your activities and interests [GDS] | 3049765 | Observation | LOINC |
| | | Is it easy for you to make decisions [GDS] | 3052959 | Observation | LOINC |
| | | Is it hard for you to get started on new projects [GDS] | 3051378 | Observation | LOINC |
| | | Is your mind as clear as it used to be [GDS] | 3052338 | Observation | LOINC |
| | | Often feel depressed | 3052936 | Observation | LOINC |
| Hamilton Anxiety Rating Scale (HAM-A) | Assessment-level | Hamilton Anxiety Rating Scale | 37157670 | Measurement | SNOMED |
| | Subscale-level | Unmapped | | | |
| | Item-level | Unmapped | | | |
| Hamilton Depression Rating Scale (HAM-D) | Assessment-level | Hamilton rating scale for depression | 4159709 | Measurement | SNOMED |
| | Item-level | Unmapped | | | |
| Interpersonal Reactivity Inventory (IRI) | Assessment-level | Unmapped | | | |
| | Subscale-level | Unmapped | | | |
| | Item-level | Unmapped | | | |
| Patient Health Questionnaire 8-item (PHQ-8) | Assessment-level | Patient Health Questionnaire 8 item (PHQ-8) [Reported] | 36660099 | Observation | LOINC |
| | | Patient Health Questionnaire 8 item (PHQ-8) total score [Reported] | 36660083 | Observation | LOINC |
| | Item-level | Feeling bad about yourself - or that you are a failure or have let yourself or your family down in last 2 weeks [Reported.PHQ] | 3043801 | Observation | LOINC |
| | | Feeling down, depressed, or hopeless in last 2 weeks | 3045858 | Observation | LOINC |
| | | Feeling tired or having little energy in last 2 weeks [Reported.PHQ] | 3044964 | Observation | LOINC |
| | | Little interest or pleasure in doing things in last 2 weeks | 3042924 | Observation | LOINC |
| | | Little interest or pleasure in doing things in last 2 weeks [Reported.PHQ] | 1260025 | Observation | LOINC |
| | | Moving or speaking so slowly that other people could have noticed. Or the opposite - being so fidgety or restless that you have been moving around a lot more than usual in last 2 weeks [Reported.PHQ] | 3043785 | Observation | LOINC |
| | | Poor appetite or overeating in last 2 weeks [Reported.PHQ] | 3044098 | Observation | LOINC |
| | | Trouble concentrating on things, such as reading the newspaper or watching television in last 2 weeks [Reported.PHQ] | 3045019 | Observation | LOINC |
| | | Trouble falling or staying asleep, or sleeping too much in last 2 weeks [Reported.PHQ] | 3045933 | Observation | LOINC |

| C. Psychosocial Assessments | | | | | |
|---|---|---|---|---|---|
| **Assessment** | **Granularity Level** | **OMOP Concept Name** | **OMOP Concept ID** | **OMOP Domain** | **Source Vocabulary** |
| Patient Health Questionnaire 9-item (PHQ-9) | Assessment-level | PHQ-9 - Patient health questionnaire 9 | 44804610 | Measurement | SNOMED |
| | | PHQ-9 quick depression assessment panel [Reported.PHQ] | 3044486 | Observation | LOINC |
| | | Patient Health Questionnaire 9 item (PHQ-9) total score [Reported] | 3042932 | Observation | LOINC |
| | Item-level | Feeling bad about yourself - or that you are a failure or have let yourself or your family down in last 2 weeks [Reported.PHQ] | 3043801 | Observation | LOINC |
| | | Feeling bad about yourself - or that you are a failure or have let yourself or your family down in last 2 weeks.frequency [Reported PHQ-9 CMS] | 40757778 | Observation | LOINC |
| | | Feeling bad about yourself - or that you are a failure or have let yourself or your family down in last 2 weeks.presence [Reported PHQ-9 CMS] | 40757777 | Observation | LOINC |
| | | Feeling down, depressed, or hopeless in last 2 weeks | 3045858 | Observation | LOINC |
| | | Feeling tired or having little energy in last 2 weeks [Reported.PHQ] | 3044964 | Observation | LOINC |
| | | Feeling tired or having little energy in last 2 weeks.frequency [Observed PHQ-9 CMS] | 40757796 | Observation | LOINC |
| | | Feeling tired or having little energy in last 2 weeks.frequency [Reported PHQ-9 CMS] | 40757774 | Observation | LOINC |
| | | Feeling tired or having little energy in last 2 weeks.presence [Observed PHQ-9 CMS] | 40757795 | Observation | LOINC |
| | | Feeling tired or having little energy in last 2 weeks.presence [Reported PHQ-9 CMS] | 40757773 | Observation | LOINC |
| | | How difficult have these made it for you to do your work, take care of things at home, or get along with other people [Reported.PHQ] | 40772146 | Observation | LOINC |
| | | Little interest or pleasure in doing things in last 2 weeks | 3042924 | Observation | LOINC |
| | | Little interest or pleasure in doing things in last 2 weeks [Reported.PHQ] | 1260025 | Observation | LOINC |
| | | Little interest or pleasure in doing things in last 2 weeks.frequency [Observed PHQ-9 CMS] | 40757790 | Observation | LOINC |
| | | Little interest or pleasure in doing things in last 2 weeks.frequency [Reported PHQ-9 CMS] | 40757768 | Observation | LOINC |
| | | Little interest or pleasure in doing things in last 2 weeks.presence [Observed PHQ-9 CMS] | 40757789 | Observation | LOINC |
| | | Little interest or pleasure in doing things in last 2 weeks.presence [Reported PHQ-9 CMS] | 40757767 | Observation | LOINC |

| C. Psychosocial Assessments | | | | | |
|---|---|---|---|---|---|
| **Assessment** | **Granularity Level** | **OMOP Concept Name** | **OMOP Concept ID** | **OMOP Domain** | **Source Vocabulary** |
| Patient Health Questionnaire 9-item (PHQ-9) *(cont.)* | Item-level *(cont.)* | Moving or speaking so slowly that other people could have noticed. Or the opposite - being so fidgety or restless that you have been moving around a lot more than usual in last 2 weeks [Reported.PHQ] | 3043785 | Observation | LOINC |
| | | Moving or speaking so slowly that other people could have noticed. Or the opposite - being so fidgety or restless that you have been moving around a lot more than usual in last 2 weeks.frequency [Reported PHQ-9 CMS] | 40757782 | Observation | LOINC |
| | | Moving or speaking so slowly that other people could have noticed. Or the opposite - being so fidgety or restless that you have been moving around a lot more than usual in last 2 weeks.presence [Reported PHQ-9 CMS] | 40757781 | Observation | LOINC |
| | | Moving or speaking so slowly that other people have noticed. Or the opposite - being so fidgety or restless that (s)he has been moving around a lot more than usual in last 2 weeks.frequency [Observed PHQ-9 CMS] | 40758034 | Observation | LOINC |
| | | Moving or speaking so slowly that other people have noticed. Or the opposite - being so fidgety or restless that (s)he has been moving around a lot more than usual in last 2 weeks.presence [Observed PHQ-9 CMS] | 40757803 | Observation | LOINC |
| | | Poor appetite or overeating in last 2 weeks [Reported.PHQ] | 3044098 | Observation | LOINC |
| | | Thoughts that you would be better off dead, or of hurting yourself in some way in last 2 weeks [Reported.PHQ] | 3043462 | Observation | LOINC |
| | | Thoughts that you would be better off dead, or of hurting yourself in some way in last 2 weeks.frequency [Reported PHQ-9 CMS] | 40757784 | Observation | LOINC |
| | | Thoughts that you would be better off dead, or of hurting yourself in some way in last 2 weeks.presence [Reported PHQ-9 CMS] | 40757783 | Observation | LOINC |
| | | Trouble concentrating on things, such as reading the newspaper or watching television in last 2 weeks [Reported.PHQ] | 3045019 | Observation | LOINC |
| | | Trouble concentrating on things, such as reading the newspaper or watching television in last 2 weeks.frequency [Observed PHQ-9 CMS] | 40757802 | Observation | LOINC |

| C. Psychosocial Assessments | | | | | |
|---|---|---|---|---|---|
| **Assessment** | **Granularity Level** | **OMOP Concept Name** | **OMOP Concept ID** | **OMOP Domain** | **Source Vocabulary** |
| Patient Health Questionnaire 9-item (PHQ-9) *(cont.)* | Item-level *(cont.)* | Trouble concentrating on things, such as reading the newspaper or watching television in last 2 weeks.frequency [Reported PHQ-9 CMS] | 40757780 | Observation | LOINC |
| | | Trouble concentrating on things, such as reading the newspaper or watching television in last 2 weeks.presence [Observed PHQ-9 CMS] | 40757801 | Observation | LOINC |
| | | Trouble concentrating on things, such as reading the newspaper or watching television in last 2 weeks.presence [Reported PHQ-9 CMS] | 40757779 | Observation | LOINC |
| | | Trouble falling or staying asleep, or sleeping too much in last 2 weeks [Reported.PHQ] | 3045933 | Observation | LOINC |
| | | Trouble falling or staying asleep, or sleeping too much in last 2 weeks.frequency [Observed PHQ-9 CMS] | 40757794 | Observation | LOINC |
| | | Trouble falling or staying asleep, or sleeping too much in last 2 weeks.frequency [Reported PHQ-9 CMS] | 40757772 | Observation | LOINC |
| | | Trouble falling or staying asleep, or sleeping too much in last 2 weeks.presence [Observed PHQ-9 CMS] | 40757793 | Observation | LOINC |
| | | Trouble falling or staying asleep, or sleeping too much in last 2 weeks.presence [Reported PHQ-9 CMS] | 40757771 | Observation | LOINC |
| Revised Social Anhedonia Scale (RSAS) | Assessment-level | Unmapped | | | |
| | Item-level | Unmapped | | | |
| Starkstein Apathy Scale (SAS) | Assessment-level | Unmapped | | | |
| | Item-level | Unmapped | | | |
| Toronto Alexithymia Scale 20-item (TAS-20) | Assessment-level | Unmapped | | | |
| | Subscale-level | Unmapped | | | |
| | Item-level | Unmapped | | | |

| D. Sensorimotor Assessments | | | | | |
|---|---|---|---|---|---|
| **Assessment** | **Granularity Level** | **OMOP Concept Name** | **OMOP Concept ID** | **OMOP Domain** | **Source Vocabulary** |
| Action Research Arm Test (ARAT) | Assessment-level | Action research arm test | 36684870 | Measurement | SNOMED |
| | Subscale-level | Unmapped | | | |
| | Item-level | Unmapped | | | |
| Activities Specific Balance Confidence Scale (ABC) | Assessment-level | Activities specific balance confidence scale | 40481019 | Measurement | SNOMED |
| | Item-level | Unmapped | | | |
| Berg Balance Scale (BBS) | Assessment-level | Berg Balance Scale | 4326493 | Measurement | SNOMED |
| | | Berg balance test | 4232747 | Measurement | SNOMED |
| | Item-level | Unmapped | | | |
| Box and Blocks Test (BBT) | Assessment-level | Unmapped | | | |

| D. Sensorimotor Assessments | | | | | |
|---|---|---|---|---|---|
| **Assessment** | **Granularity Level** | **OMOP Concept Name** | **OMOP Concept ID** | **OMOP Domain** | **Source Vocabulary** |
| Compound Motor Evoked Potential (cMEP) | Assessment-level | Unmapped | | | |
| Dynamic Gait Index (DGI) | Assessment-level | Dynamic gait index | 40481604 | Measurement | SNOMED |
| | Item-level | Unmapped | | | |
| Edinburgh Handedness Inventory (EHI) | Assessment-level | Unmapped | | | |
| | Item-level | Unmapped | | | |
| Finger Tapping Test (FTT) | Assessment-level | Finger tapping test | 4159700 | Measurement | SNOMED |
| Five Times Sit to Stand Test (FTSST) | Assessment-level | Unmapped | | | |
| Fugl-Meyer Upper Extremity Motor Assessment (FMUE) | Assessment-level | Unmapped | | | |
| | Subscale-level | Unmapped | | | |
| | Item-level | Unmapped | | | |
| Fugl-Meyer Lower Extremity Motor Assessment (FMLE) | Assessment-level | Unmapped | | | |
| | Subscale-level | Unmapped | | | |
| | Item-level | Unmapped | | | |
| Functional Ambulation Category (FAC) | Assessment-level | Unmapped | | | |
| Functional Gait Assessment (FGA) | Assessment-level | Unmapped | | | |
| | Item-level | Unmapped | | | |
| Grip Strength (Dynamometer) | Assessment-level | Dynamometer pinch and gross grip scale | 44802591 | Measurement | SNOMED |
| | | Grip Strength Test [NIH Toolbox] | 42529268 | Measurement | LOINC |
| | | Grip strength | 4089159 | Measurement | SNOMED |
| | | Grip strength Hand - left Dynamometer | 42529367 | Measurement | LOINC |
| | | Grip strength Hand - right Dynamometer | 42529368 | Measurement | LOINC |
| | | Grip strength of left hand | 44805437 | Measurement | SNOMED |
| | | Grip strength of right hand | 44805438 | Measurement | SNOMED |
| Grooved Pegboard Test | Assessment-level | Unmapped | | | |
| Jebsen Taylor Hand Function Test (JTHFT) | Assessment-level | Jebsen hand function test | 4169153 | Measurement | SNOMED |
| | Item-level | Unmapped | | | |
| Medical Research Council Scale for Muscle Strength | Assessment-level | MRC (Medical Research Council) Muscle scale | 35610373 | Measurement | SNOMED |
| | | MRC grade - muscle power | 4093203 | Measurement | SNOMED |
| | | MRC grade - muscle power - finding | 4272225 | Condition | SNOMED |
| | | Medical Research Council motor power scale | 4169171 | Measurement | SNOMED |
| Modified Ashworth Scale (MAS) | Assessment-level | Modified Ashworth Scale | 35623426 | Measurement | SNOMED |
| Motor Activity Log (MAL) | Assessment-level | Unmapped | | | |
| | Subscale-level | Unmapped | | | |
| | Item-level | Unmapped | | | |
| Motor Evoked Potential (MEP) | Assessment-level | Motor evoked potentials monitoring | 4266033 | Observation | SNOMED |
| Motricity Index (MI) | Assessment-level | Motricity index | 4169181 | Measurement | SNOMED |
| | Subscale-level | Unmapped | | | |
| | Item-level | Unmapped | | | |

| D. Sensorimotor Assessments | | | | | |
|---|---|---|---|---|---|
| **Assessment** | **Granularity Level** | **OMOP Concept Name** | **OMOP Concept ID** | **OMOP Domain** | **Source Vocabulary** |
| Nine Hole Peg Test (NHPT) | Assessment-level | 9-Hole Pegboard Dexterity Test [NIH Toolbox] | 42528699 | Measurement | LOINC |
| | | Nine hole peg test | 4165291 | Measurement | SNOMED |
| | Item-level | 9-Hole Pegboard Dexterity - unadjusted scale score Hand.dominant [NIH Toolbox] | 42528541 | Measurement | LOINC |
| | | 9-Hole Pegboard Dexterity Test Time Hand - left [NIH Toolbox] | 42529303 | Measurement | LOINC |
| | | 9-Hole Pegboard Dexterity Test Time Hand - right [NIH Toolbox] | 42529304 | Measurement | LOINC |
| Over-ground Self-selected Walking Speeds (SSWS) | Assessment-level | Unmapped | | | |
| Pinch Strength (Pinch Gauge/Dynamometer) | Assessment-level | Dynamometer pinch and gross grip scale | 44802591 | Measurement | SNOMED |
| Purdue Pegboard Test | Assessment-level | Purdue pegboard scale | 4158641 | Measurement | SNOMED |
| | | Purdue pegboard test | 4224278 | Procedure | SNOMED |
| | Item-level | Unmapped | | | |
| Resistance to Passive Movement Scale (REPAS) | Assessment-level | Unmapped | | | |
| | Subscale-level | Unmapped | | | |
| | Item-level | Unmapped | | | |
| Rivermead Mobility Index (RMI) | Assessment-level | Rivermead Mobility Index | 44811906 | Measurement | SNOMED |
| | Item-level | Unmapped | | | |
| Rivermead Motor Assessment (RMA) | Assessment-level | Rivermead motor assessment | 4158744 | Measurement | SNOMED |
| | Subscale-level | Unmapped | | | |
| | Item-level | Unmapped | | | |
| Six Minute Walk Test (6MWT) | Assessment-level | 6 minute walk test distance | 606289 | Measurement | SNOMED |
| | | 6-minute walk test | 4099101 | Procedure | SNOMED |
| | | Six minute walk test | 40766814 | Measurement | LOINC |
| Ten Meter Walk Test (10MWT) | Assessment-level | 10-meter walking distance [Time] | 1259995 | Observation | LOINC |
| | | Timed 10 meter walk | 4197638 | Procedure | SNOMED |
| | Item-level | Unmapped | | | |
| Two Point Discrimination Test | Assessment-level | Two point discrimination distance | 4114296 | Observation | SNOMED |
| | | Two point static discrimination - finding | 4270716 | Condition | SNOMED |
| | | Two point static discrimination response | 4151499 | Observation | SNOMED |
| Wolf Motor Function Test (WMFT) | Assessment-level | Unmapped | | | |
| | Subscale-level | Unmapped | | | |
| | Item-level | Unmapped | | | |
| von Frey Hair Test (VFHT) / Monofilament Test | Assessment-level | Monofilament foot sensation test | 4047085 | Procedure | SNOMED |

### SUPPLEMENTAL FIGURES

**Supplemental Figure 1.** Number of Observational Medical Outcomes Partnership (OMOP) standard concepts and domains for each mapped demographics and medical history (DMH) data element (n = 39)

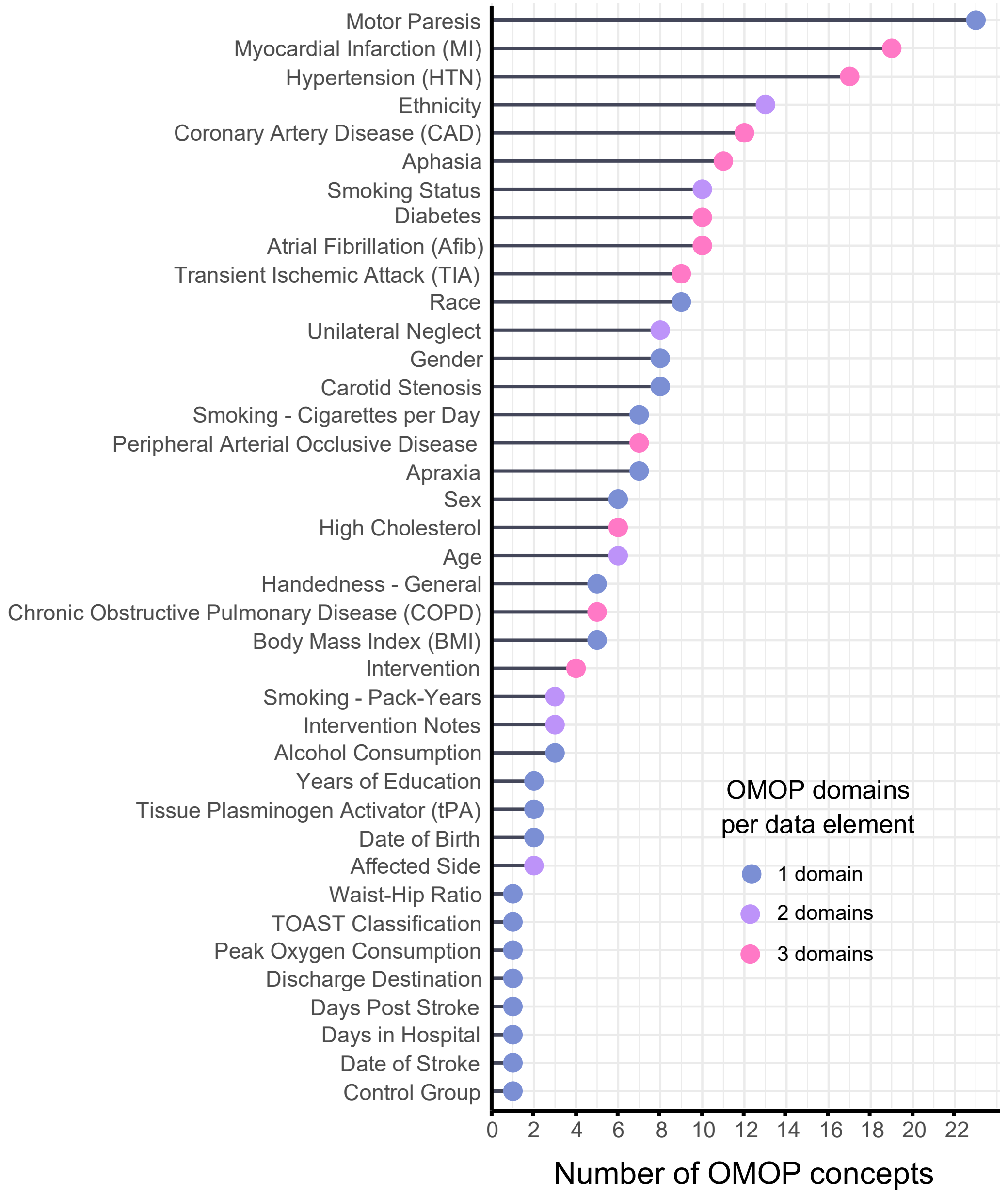

**Supplemental Figure 2.** Number of Observational Medical Outcomes Partnership (OMOP) standard concepts and domains for each mapped assessment-level data element (n = 54)

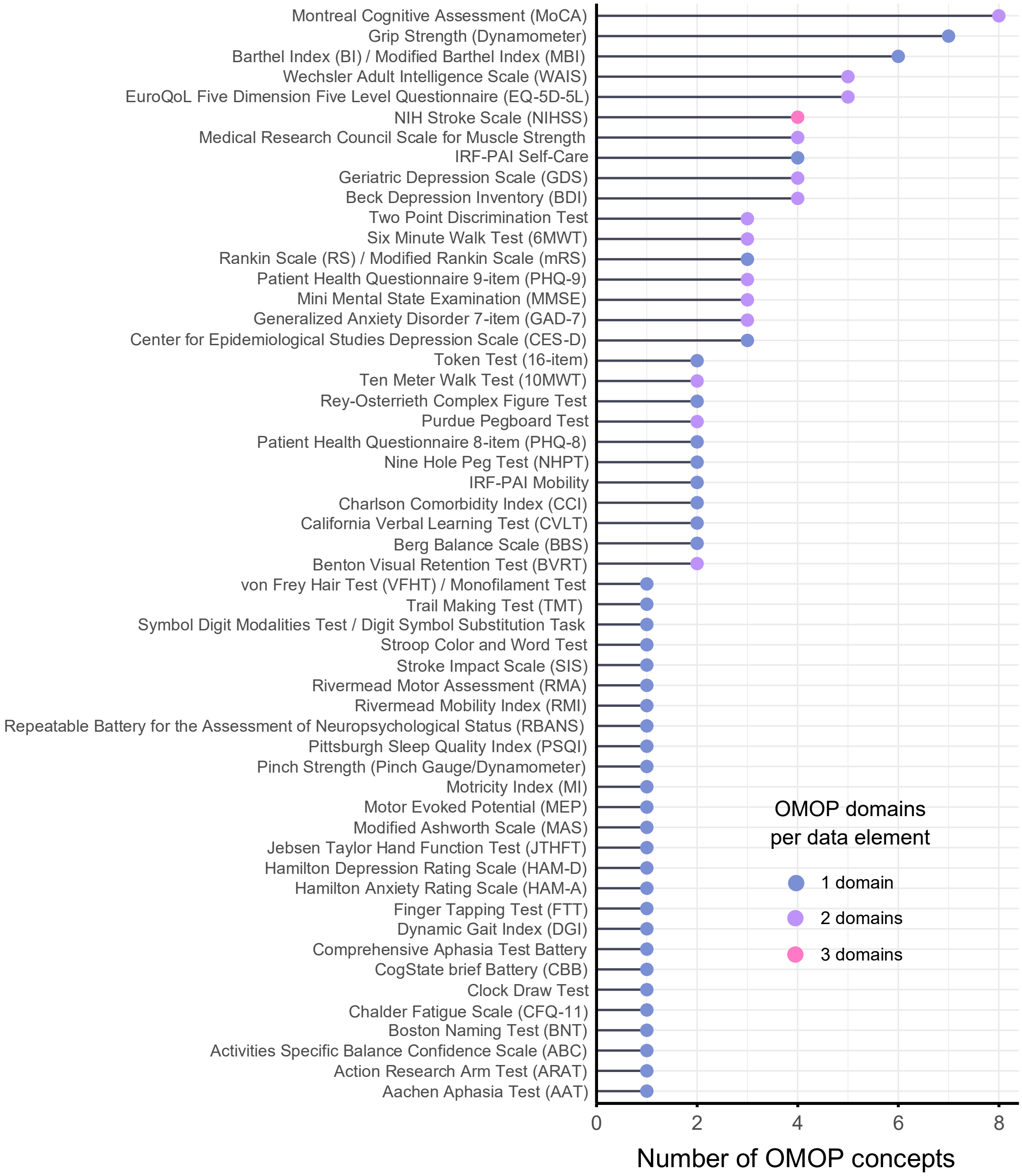
